# Single-Cell Analysis Reveals Inflammatory–Immunosuppressive Niches Associated with Suboptimal Response to Daratumumab-Based Therapy in AL Amyloidosis

**DOI:** 10.64898/2026.03.28.26349317

**Authors:** Xuezhu Wang, Xinyi Xiong, Chengyang Xu, Hongxiao Han, Ai Guan, Yajuan Gao, Qi Yan, Kaini Shen, Jian Li

## Abstract

Systemic light-chain amyloidosis is caused by plasma cell clones that secrete misfolded free light chains. Although daratumumab is used for first-line therapy, mechanisms underlying suboptimal response remain unclear. We generated a bone marrow single-cell atlas from 30 patients treated with a first-line daratumumab-based regimen. Suboptimal responders were defined as not achieving a very good partial response within six cycles. In the t(11;14) subgroup, amyloidogenic plasma cells from suboptimal responders exhibited lower baseline protein-translation and cell-cell adhesion programs than those from good responders. The bone marrow of suboptimal responders exhibited increased baseline (i) prostaglandin E_2_-EP2/EP4 signaling with myeloid-derived suppressor cell–like CD14⁺ monocytes expressing *PTGS2*; and (ii) non-classical MHC class I signaling between plasma cells and the microenvironment. Following treatment, myeloid and NK-cell dysfunction, T-cell exhaustion, and IFN-γ responses were more pronounced in suboptimal responders than in good responders. This atlas reveals inflammatory–immunosuppressive bone marrow niches that underpinned suboptimal response.

## INTRODUCTION

Systemic light-chain amyloidosis (AL) is a systemic disorder in which slowly proliferating plasma cell (PC) clones secrete misfolded immunoglobulin light chains. These amyloidogenic light chains aggregate into fibrils that deposit in vital organs, particularly the heart and kidney, leading to progressive organ dysfunction. From a hematologic standpoint, the primary therapeutic objective is to eradicate the amyloidogenic PC clone and thereby suppress the production of pathogenic light chains, and thus inhibit their deposition in organs, with rapid suppression of free light chains being essential to prevent further injury and to salvage organ function whenever possible^1^.

Daratumumab, an anti-CD38 monoclonal antibody that mainly targets PCs, has been established as a key component of the first-line therapy for AL by the ANDROMEDA trial since 2021^2^. However, approximately 20% of patients treated with daratumumab-based therapy fail to achieve very good partial response (VGPR)^3^. Given daratumumab’s central role as frontline therapy for AL, baseline biomarkers that predict hematologic response are crucial for identifying individuals unlikely to benefit, so that they can be redirected to alternative treatments before therapy begins. Chakraborty et al. identified 1q21 gain as a statistically significant predictor of inferior hematologic response and event-free survival in a 283-patient cohort^4^. However, because ∼80% of patients do not harbor 1q21 gain, there remains a clear need for biomarkers and mechanisms that are broadly applicable to patients with AL.

Daratumumab’s mechanism of action in plasma cell disorders is well studied, yet the specific immune processes driving resistance, particularly in AL, remain under investigation. Conceptually, resistance should arise from disruptions or suppression in the following immune pathways through which daratumumab exerts its effects. Daratumumab binds CD38 on PCs and CD38 immune cells in the microenvironment, including natural killer (NK) cells^5^ and myeloid-derived suppressor cells (MDSCs)^6^, and then induces their depletion through several immune effector pathways^7^. These mainly include (i) antibody-dependent cellular cytotoxicity (ADCC), in which engagement of Fcγ receptors on NK cells initiates cytotoxicity; (ii) antibody-dependent cellular phagocytosis (ADCP), mediated by macrophages or monocytes that recognize and engulf daratumumab-opsonized PCs via Fcγ receptor binding; and (iii) complement-dependent cytotoxicity (CDC), in which complement components bind to daratumumab and activate the complement cascade^8^.

In patients who are resistant to daratumumab-based therapy, these immune responses are disrupted or suppressed by an immunosuppressive bone marrow (BM) niche. Inflammation is a plausible contributing factor to this kind of niche, given that it is a recognized feature of the BM microenvironment in AL. The most compelling recent evidence comes from Gort-Freitas et al., who used single-cell RNA sequencing (scRNA-seq) to compare BM cells in patients with AL versus age-matched healthy donors. They observed elevated interferon (IFN)-γ responses across multiple immune lineages and an expansion of non-traditional CD16⁺ monocytes^9^. These findings are echoed by earlier reports that the BM microenvironment in AL is inherently inflammatory^10^, likely associated with the toxicity of amyloidogenic light chains. Together, these findings support the presence of an inflammatory BM niche that may be permissive to amyloidogenic PCs while impairing immune effector function.

Based on these findings, we hypothesize that resistance to daratumumab is associated with two interrelated processes: (i) intrinsic gene expression programs (GEPs) of amyloidogenic PC clones^11^, and (ii) inflammatory and immunosuppressive dysregulation across multiple immune cell types within the BM niche. To test these hypotheses, we analyzed scRNA-seq samples from a prospective cohort of patients with AL treated with a daratumumab-based regimen. Through this study, we aim to define the mechanisms that are associated with daratumumab resistance in AL and to provide a profile of candidates for baseline predictive biomarkers and therapeutic targets to overcome this resistance.

## RESULTS

### Single-cell profiling characterized clonal expansion of plasma cells secreting amyloidogenic light chains in AL treated with daratumumab

For the single-cell study, we recruited 30 patients with newly diagnosed AL who all received first-line daratumumab-bortezomib-dexamethasone (Dara-Vd), following the same treatment regimen and follow-up protocol as described in the previous report^12^. Good responders were defined as achieving ≥VGPR by hematologic response criteria, whereas suboptimal responders were defined as achieving <VGPR within six cycles of Dara-Vd (Fig. 1a).

**Fig. 1.**
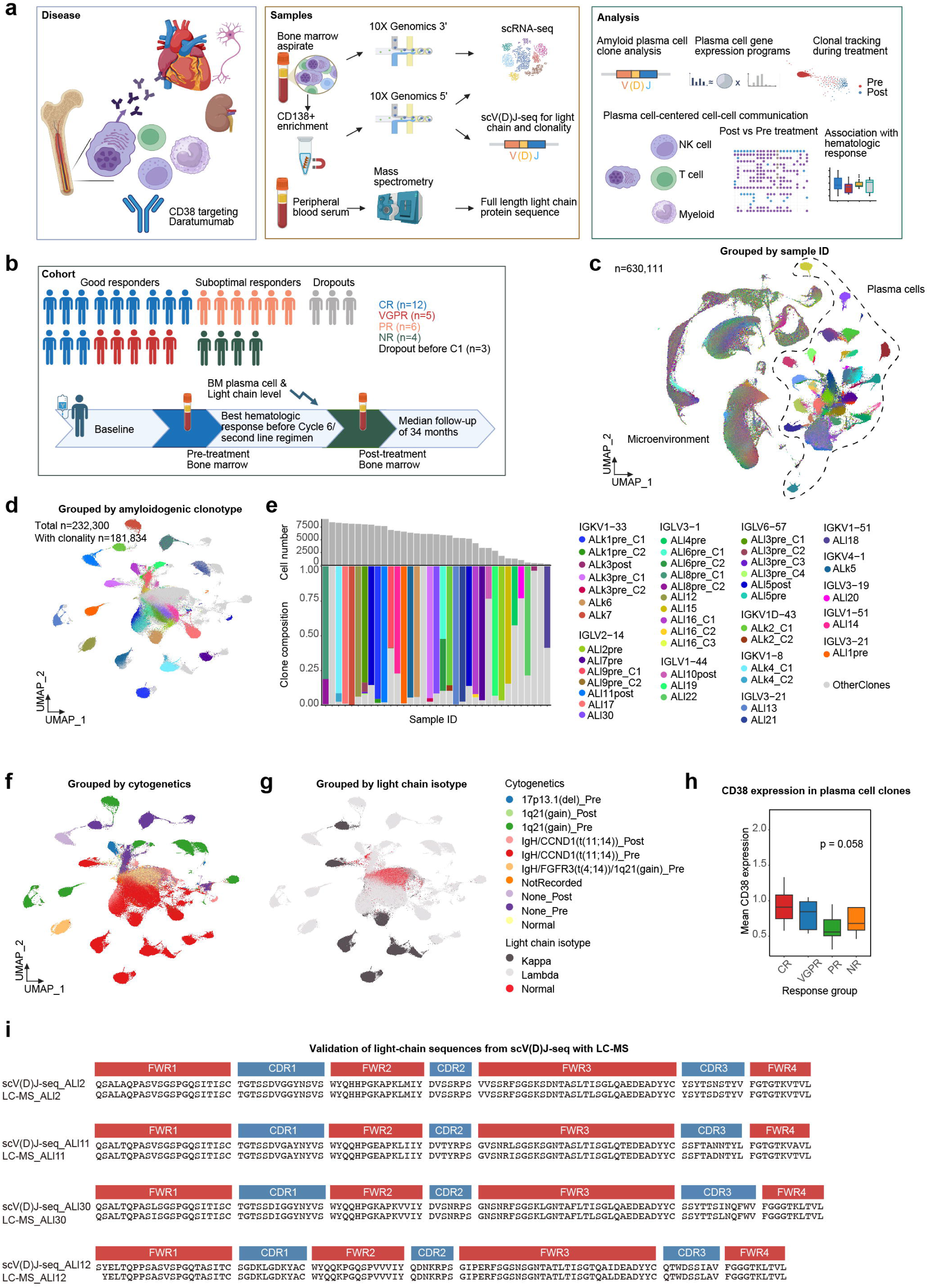
Bone marrow cell atlas of systemic light-chain amyloidosis treated with a first-line daratumumab-based regimen. **(a)** Flowchart of the study design and analysis strategy. scRNA-seq: single-cell RNA sequencing; scV(D)J-seq: single-cell V(D)J sequencing. **(b)** Cohort design and patient outcome. CR: Complete response; VGPR: Very good partial response; PR: Partial response; NR: No response. **(c)** Uniform manifold approximation and projection (UMAP) visualization of bone marrow (BM) plasma cells (PCs) and microenvironment. **(d)** UMAP visualization of predominant PC clones determined by scV(D)J-seq and scRNA-seq. **(e)** Bar plot and stack plot showing PC number and clone composition in each sample. **(f)** UMAP visualization of PCs, grouped and colored by cytogenetic abnormalities. **(g)** UMAP visualization of PCs, grouped and colored by light-chain isotype. **(h)** CD38 expression in amyloidogenic PC clones, grouped by hematologic response to daratumumab-bortezomib-dexamethasone (Dara-Vd). **(i)** Validation of full-length light-chain sequences generated by scV(D)J-seq, using liquid chromatography mass spectrometry (LC–MS).

At the time of enrollment, 7/30 (23.3%) patients had κ light chain AL, 29/30 (96.7%) had cardiac involvement, 7/30 (23.3%) had renal involvement, and 28/30 (93.3%) were Mayo 2004 stage I to IIIa. Common cytogenetic features included t(11;14) in 14/30 (46.7%) patients and 1q21 gain in 6/30 (20.0%). Three patients experienced early cardiac events before response evaluation in Cycle 1 and therefore dropped out of the study (Fig. 1b). Among the remaining 27 patients, 10/27 (37.0%) showed a suboptimal hematologic response within six cycles of Dara-Vd before switching to the next-line therapy, followed by a median follow-up of 34 months (by December 2025). Regarding sample collection, 17 were good responders (7 with paired baseline and post-treatment samples) and 10 were suboptimal responders (4 with paired samples, 2 with post-treatment samples only) (Fig. 1b; Table 1; Supplementary Table S1). In addition, two healthy volunteers with normal BM served as controls, providing polyclonal PCs.

**Table 1.**
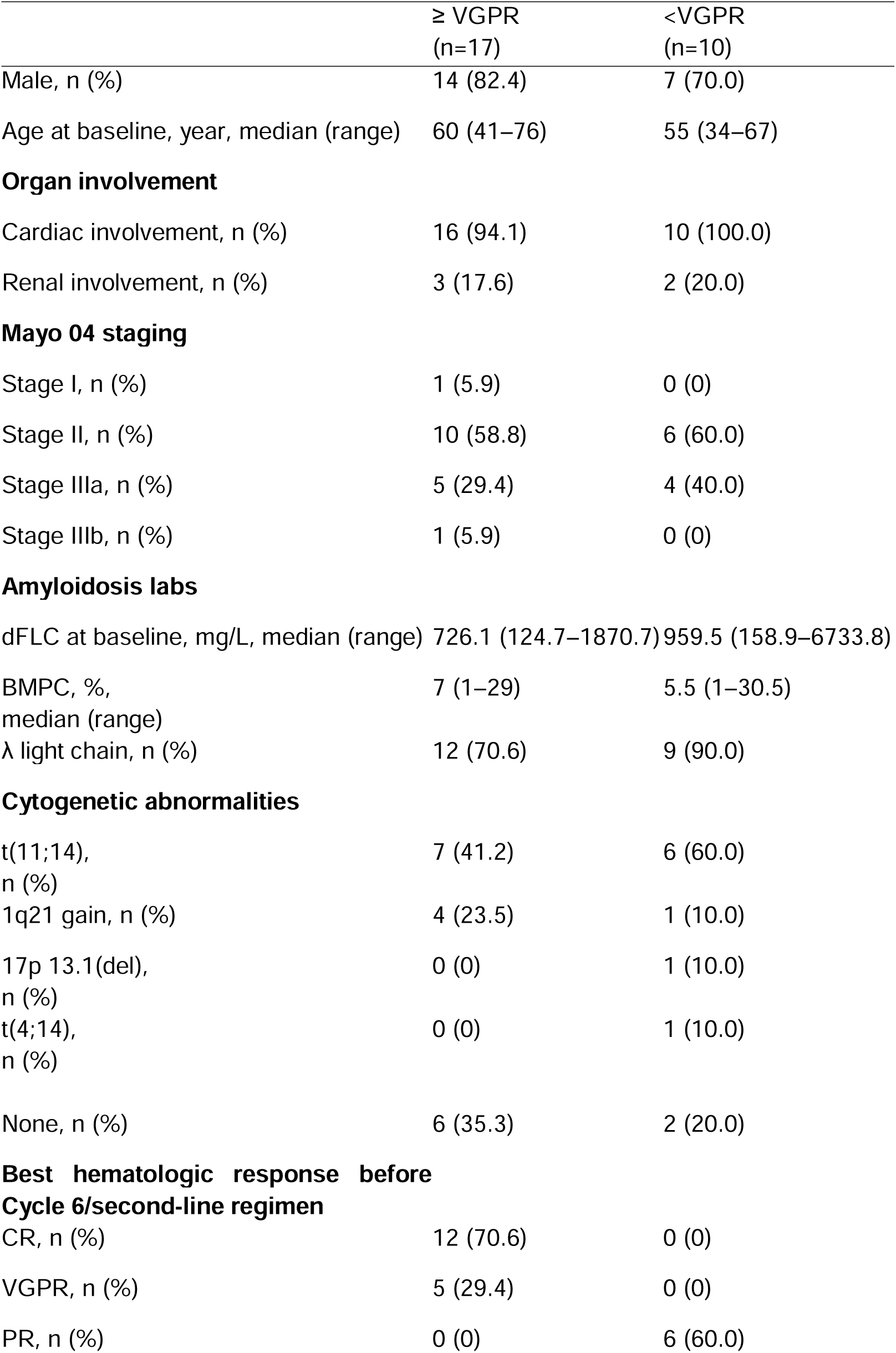
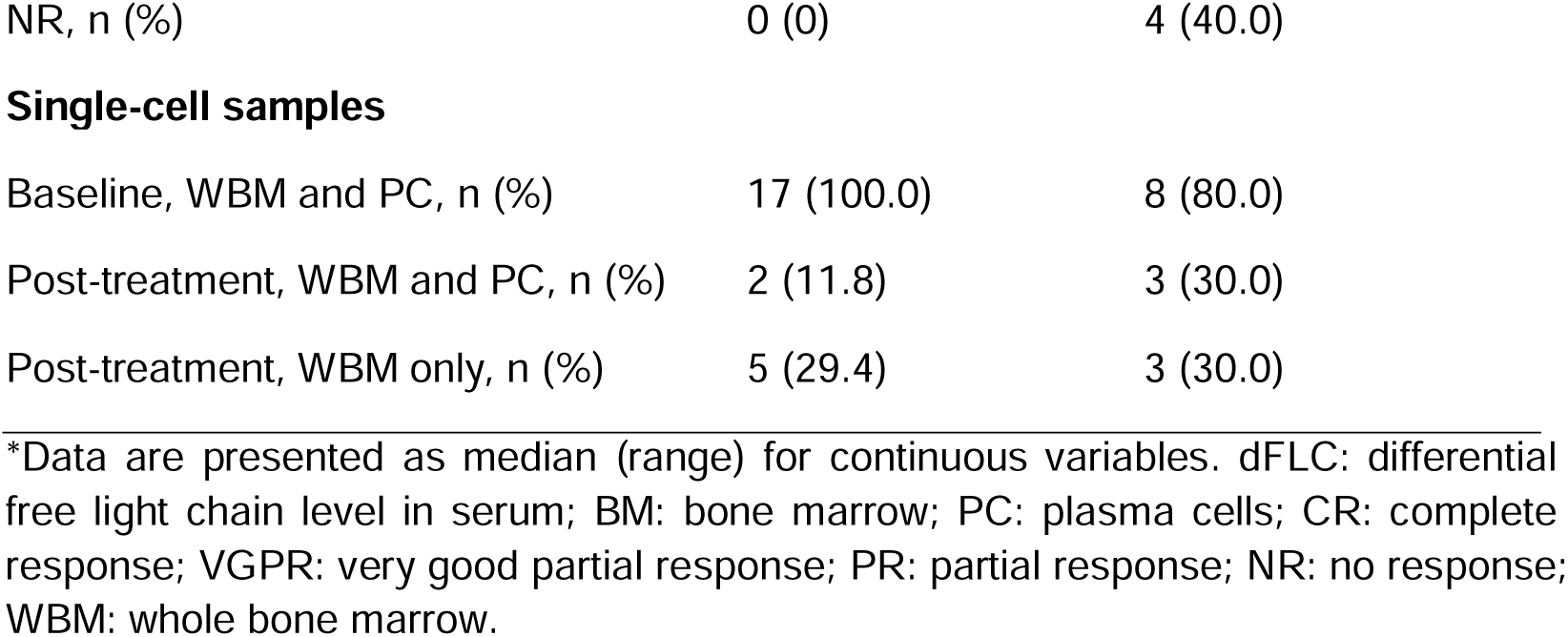
Clinical characteristics of patients in the single-cell cohort.

|  | ≥ VGPR<br>(n=17) | <VGPR<br>(n=10) |
| --- | --- | --- |
| Male, n (%) | 14 (82.4) | 7 (70.0) |
| Age at baseline, year, median (range) | 60 (41–76) | 55 (34–67) |
| **Organ involvement** |  |  |
| Cardiac involvement, n (%) | 16 (94.1) | 10 (100.0) |
| Renal involvement, n (%) | 3 (17.6) | 2 (20.0) |
| **Mayo 04 staging** |  |  |
| Stage I, n (%) | 1 (5.9) | 0 (0) |
| Stage II, n (%) | 10 (58.8) | 6 (60.0) |
| Stage IIIa, n (%) | 5 (29.4) | 4 (40.0) |
| Stage IIIb, n (%) | 1 (5.9) | 0 (0) |
| **Amyloidosis labs** |  |  |
| dFLC at baseline, mg/L, median (range) | 726.1 (124.7–1870.7) | 959.5 (158.9–6733.8) |
| BMPC, %, median (range) | 7 (1–29) | 5.5 (1–30.5) |
| λ light chain, n (%) | 12 (70.6) | 9 (90.0) |
| **Cytogenetic abnormalities** |  |  |
| t(11;14), n (%) | 7 (41.2) | 6 (60.0) |
| 1q21 gain, n (%) | 4 (23.5) | 1 (10.0) |
| 17p 13.1(del), n (%) | 0 (0) | 1 (10.0) |
| t(4;14), n (%) | 0 (0) | 1 (10.0) |
| None, n (%) | 6 (35.3) | 2 (20.0) |
| **Best hematologic response before Cycle 6/second-line regimen** |  |  |
| CR, n (%) | 12 (70.6) | 0 (0) |
| VGPR, n (%) | 5 (29.4) | 0 (0) |
| PR, n (%) | 0 (0) | 6 (60.0) |
| NR, n (%) | 0 (0) | 4 (40.0) |
| **Single-cell samples** |  |  |
| Baseline, WBM and PC, n (%) | 17 (100.0) | 8 (80.0) |
| Post-treatment, WBM and PC, n (%) | 2 (11.8) | 3 (30.0) |
| Post-treatment, WBM only, n (%) | 5 (29.4) | 3 (30.0) |

\*Data are presented as median (range) for continuous variables. dFLC: differential free light chain level in serum; BM: bone marrow; PC: plasma cells; CR: complete response; VGPR: very good partial response; PR: partial response; NR: no response; WBM: whole bone marrow.

From this cohort, we generated a BM atlas comprising 630,111 single cells from 78 scRNA-seq samples, including PCs, myeloid lineage, T cell lineage, NK cell lineage, and B cell lineage (Fig. 1c) after filtering cells with low gene number/unique molecular identifier (UMI) counts, high mitochondrial fraction/hemoglobin fraction (Supplementary Fig. S1a), or doublets. A total of 232,300 PCs were distinguished from other cell types based on high *CD38*, *SDC1* (*CD138*), *XBP1,* and *PRDM1* expression, and high immunoglobulin expression among all reads (Supplementary Fig. S1b). Using single-cell V(D)J sequencing (scV(D)J-seq) to define clonotypes from B-cell receptor (BCR) sequences (Fig. 1d), we found that the dominant PC clone was markedly expanded in AL, accounting for an average of 79.3% of PCs in baseline samples. Patient IDs comprise the amyloidogenic light-chain isotype (“k” or “l”), and the ID number (e.g., “ALk1”). IGKV/IGLV labels denote the clone’s immunoglobulin variable region gene. Clones sharing the same isotype but differing in V(D)J clonotype due to subtle light-chain sequence variations were designated by clone size (e.g., “C1”, “C2”) (Fig. 1e).

Consistent with prior studies of plasma cell disorders (including multiple myeloma, MM), amyloidogenic plasma cells exhibited substantial inter-patient transcriptomic heterogeneity. This variation was associated primarily with cytogenetic abnormalities, which are established drivers of plasma cell states, and with immunoglobulin light-chain isotype, a feature that may be particularly relevant to AL (Fig. 1f–g). PC clones highly expressed genes commonly upregulated in plasma cell diseases and associated with specific cytogenetic abnormalities^13,14^ (Supplementary Fig. S1c). In line with previous reports, patients with suboptimal hematologic responses tended to have lower *CD38* expression on PC clones (p = 0.058), suggesting reduced availability of the binding targets of daratumumab (Fig. 1h)^15^.

We validated that the expanded PC clones are amyloidogenic with the following evidence: (i) The PC clones sharing the same clonotype exhibited divergent transcriptomic states from the normal states (Supplementary Fig. S2a, S2b); (ii) Each clone expressed an exclusive λ or κ light-chain isotype, concordant with the serum light chain isotype (Supplementary Fig. S2c); and (iii) full-length light-chain sequences spanning framework region 1 (FWR1), complementarity-determining region 1 (CDR1), framework region 2 (FWR2), complementarity-determining region 2 (CDR2), framework region 3 (FWR3), complementarity-determining region 3 (CDR3), and framework region 4 (FWR4) were predicted by AlphaFold to adopt β-sheet–rich folds with high confidence (pLDDT > 90). As orthogonal validation, in four randomly selected patients, the amyloidogenic light-chain sequences identified by scV(D)J-seq in plasma cells were almost identical to those detected in corresponding serum proteins by liquid chromatography–mass spectrometry (LC–MS), a proteomic approach analogous to EasyM (Fig. 1i)^16^.

Overall, we established a single-cell prospective cohort of patients with AL receiving Dara-Vd and collected BM samples enriched in PCs at baseline and after therapy. By integrating gene expression from scRNA-seq with light-chain sequencing from scV(D)J-seq and LC–MS, we could identify the amyloidogenic PC clones at the single-cell level, which are the source of AL pathogenesis and will be the focus of our subsequent analyses.

### Convergent plasma cell gene expression programs link protein translation and endoplasmic reticulum stress responses related to amyloidogenesis

Despite substantial inter-patient heterogeneity, we sought to define GEPs shared by amyloidogenic PCs in multiple clones. To prioritize genes that are consistently differentially expressed across patients, we compared amyloidogenic PC clones with coexisting polyclonal PCs within each patient with AL. This approach extracted amyloidosis-associated differentially expressed genes (DEGs) that are robust to inter-patient heterogeneity. Compared to polyclonal PCs, amyloidogenic PCs showed coordinated upregulation of genes enriched for protein translation and ribosome biogenesis (for example, peptide-chain elongation and cytoplasmic translation), MHC class I antigen-processing and presentation, and pathways previously linked to amyloid biology, including macromolecule biosynthesis and amyloid fiber formation (Fig. 2a). We observed multiple gene signatures from various disease databases enriched for genes encoding proteins previously implicated in non-AL amyloidosis, including *CST3*, *ITM2B*, and *B2M*. These signatures suggest that the amyloidogenicity of light chains in AL is coupled to cellular processes shared with other forms of amyloidosis. In contrast to the upregulated functions, amyloid clones exhibited reduced expression of MHC class II components, B-cell activation and differentiation programs, and BCR signaling and immunoglobulin synthesis, indicating a loss of conventional B-cell/antibody functions in favor of a more amyloidogenesis-active, MHC class I–skewed state.

**Fig. 2.**
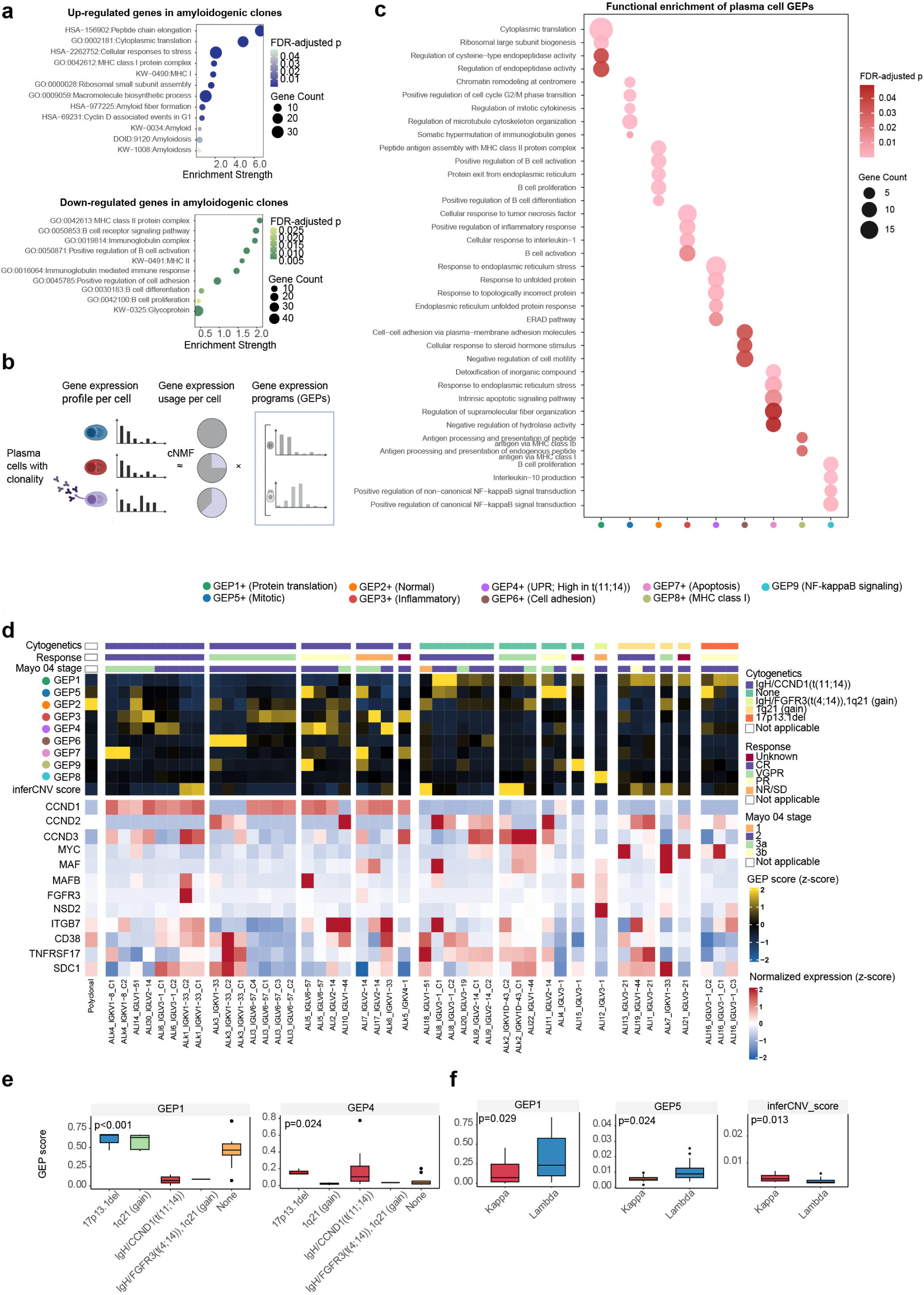
Gene expression programs in the pathogenic amyloidogenic plasma cells. **(a)** Functional enrichment of the upregulated/downregulated genes in amyloidogenic plasma cell (PC) clones compared with the polyclonal normal PCs. HSA: KEGG Human Pathway IDs; GO: Gene Ontology IDs; KW: UniPort KeyWords IDs; DOID: Disease Ontology IDs. **(b)** Consensus non-negative matrix factorization (cNMF) algorithm to infer gene expression programs (GEPs) from amyloidogenic/normal PC clones. **(c)** GO enrichment of the GEPs in amyloidogenic PC clones compared with the polyclonal normal PCs. **(d)** Heatmap showing GEP scores, inferred copy number variations (inferCNV) score, and marker gene expression across amyloidogenic PC clones, annotated with clinical features. **(e)** Differential GEP scores by cytogenetic subgroups. **(f)** Differential GEP scores and inferred copy number variations (inferCNV) scores by light-chain isotypes.

To assess the external generalizability of these amyloidogenic PC programs, we analyzed the dataset with paired scRNA-seq and scV(D)J-seq, which enabled the distinction of amyloidogenic PC clones from coexisting polyclonal PCs9 Overall, signatures upregulated in amyloidogenic PCs were similarly enriched in amyloidogenic clones in the external dataset relative to either polyclonal PCs from the same patients or polyclonal PCs from healthy donors. Conversely, signatures downregulated in amyloidogenic PC clones were consistently reduced across both comparisons. The only exception was the MHC class II program, which was modestly increased in amyloidogenic PC clones relative to healthy-donor PCs but reduced relative to within-patient polyclonal PCs (Supplementary Fig. S3a).

Following differential gene expression analysis, we applied consensus non-negative matrix factorization (cNMF) to define reference PC GEPs and to characterize differences between amyloidogenic plasma cell clones and polyclonal plasma cells (Fig. 2b; Supplementary Fig. S3b). The resulting GEPs provided a reference that we later projected onto PCs without assigned clonotypes, including minimal residual disease PCs and samples profiled by bulk RNA-seq. cNMF resolved nine biologically interpretable GEPs that are variably active across PC clones and patients. GEP1 captured protein translation pathways. GEP2 reflected B-cell activation and differentiation programs. GEP3 was related to cytokine responses, including tumor necrosis factor and interleukin-1 signaling. GEP4 included endoplasmic reticulum (ER) stress and the unfolded protein response (UPR). GEP5 was enriched for mitotic and cell-cycle regulators. GEP6 comprised cell–cell adhesion pathways. GEP7 was associated with the intrinsic apoptosis pathway and detoxification. GEP8 was related to MHC class I antigen processing and presentation. GEP9 was enriched for interleukin-10 production and NF-κB signal transduction (Fig. 2c; Supplementary Table S2).

When linked to clinical data, GEP2 was found almost exclusively active in polyclonal PCs, including those from the disease samples and healthy controls. Several genes with established mechanistic links to cytogenetic abnormalities in plasma cell diseases showed the expected expression patterns, providing an internal validation of our analytical framework. For example, high *CCND1* expression was strongly associated with t(11;14), whereas elevated *MYC* expression was associated with 1q21 gain, consistent with their known cytogenetic correlation. These established cytogenetic–transcriptome relationships further supported our newly identified associations between GEP activity and specific cytogenetic subgroups (Fig. 2d).

GEP1 activity was higher in the amyloidogenic PC clones in patients with 1q21 gain, whereas GEP4 was preferentially active in patients with t(11;14). Given that t(11;14) and t(4;14) are immunoglobulin heavy chain translocations that define biologically distinct subtypes of plasma cell disorders, these patterns suggest that cytogenetic background may shape how amyloidogenic PCs balance protein synthesis and ER-stress responses (Fig. 2e). GEP1 and GEP5 were expressed at higher levels in λ light chain AL compared to κ light chain AL, while the inferred copy number variations (inferCNV) score was higher in κ light chain AL (Fig. 2f).

Overall, our analysis delineates convergent PC GEPs that link amyloidogenesis to protein translation activity, mitotic cell-cycle regulation, and ER stress, distinguishing amyloidogenic PCs from normal PCs. These programs provide a reference framework for defining the functional states of amyloid versus polyclonal PCs, which will be used in the subsequent response-correlation analyses.

### Baseline plasma cell gene expression program predicts suboptimal response and mitotic plasma cell programs mark residual clonal plasma cells

With these PC GEPs in hand, we next asked whether their activity in amyloidogenic PCs was associated with treatment exposure and hematologic response. We used non-negative least squares (NNLS) regression to infer the GEP states of all PCs, including those from whole BM samples without scV(D)J-seq, including minimal residual disease PCs, resulting in 99.996% of PCs with inferred GEP states (Fig. 3a-b). NNLS-derived activities in PCs with clonotypes showed high concordance with the original cNMF-based inference (Supplementary Fig. S4a).

**Fig. 3.**
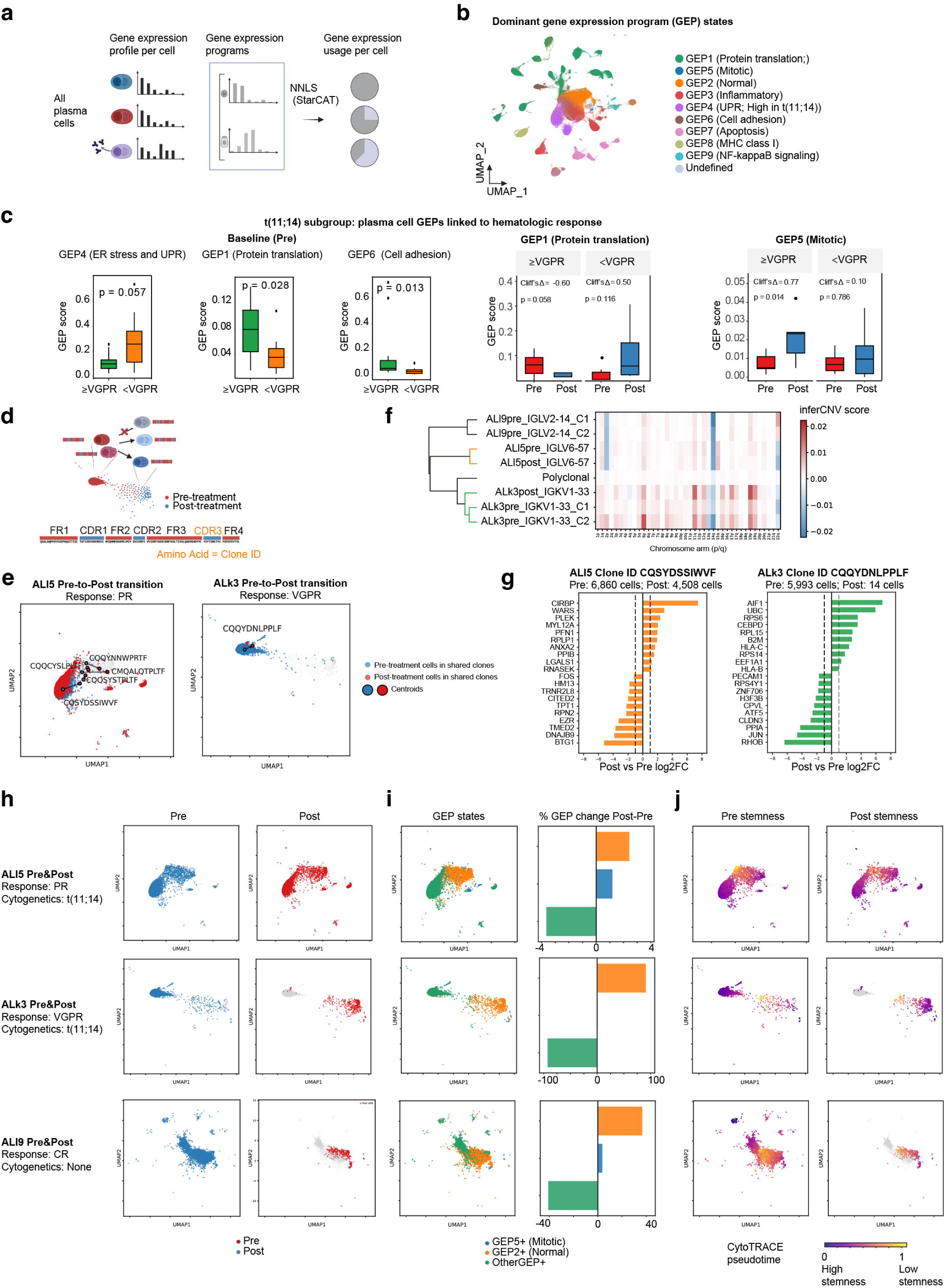
Dynamics of plasma cell clonal gene programs before and after treatment. **(a)** Non-negative least squares (NNLS) algorithm to project gene expression programs (GEPs) onto all plasma cells (PCs), including minimal residual disease PCs. **(b)** Uniform manifold approximation and projection (UMAP) visualization of dominant PC states defined by NNLS-inferred GEPs across all PCs. **(c)** Baseline and treatment-associated PC GEP scores linked to hematologic response among patients with t(11;14). **(d)** Clonal tracing with B-cell receptor (BCR) sequence to estimate the transition of PC states within clones from pre- to post-treatment. **(e)** Clone dynamics from pre- to post-treatment tracked with BCR sequence in patients showing good (VGPR) or suboptimal (PR) responses. **(f)** Inferred copy number variations (inferCNV) in amyloidogenic PC clones from pre- and post-treatment samples, clustered by Pearson correlation coefficient (r). **(g)** Bar plots showing the top upregulated and downregulated genes during the pre- to post-treatment transition within individual clones. **(h)** UMAP visualization of PCs annotated by sampling time. **(i)** UMAP visualization of GEP states, with bar plots showing changes in the fraction of cells assigned to each GEP state from pre- to post-treatment. **(j)** UMAP visualization of CytoTRACE pseudotime scores, representing inferred stemness before and after treatment.

We next used the pre- and post-treatment samples to examine how the activity of these GEPs changes over time and how it relates to hematologic response to Dara-Vd. Recent studies have identified 1q21 gain as an independent risk factor for resistance to daratumumab-based therapy. This raises an important question of how to further stratify patients without 1q21 gain, particularly those with t(11;14), who account for approximately half of patients with AL. Among patients with t(11;14), those who subsequently had suboptimal hematologic responses showed lower activity of protein translation (GEP1) and cell-cell adhesion programs (GEP6) but higher activity of ER stress programs (GEP4). During treatment, the mean activity score of the protein-translation program (GEP1) in amyloidogenic PCs increased in suboptimal responders but decreased in good responders (Fig. 3c), which was the program preferentially expressed at higher levels in patients with 1q21 gain (Fig. 3e). The mitotic programs (GEP5) and normal PC–associated programs (GEP2) became progressively more active after treatment (Fig. 3c). In an independent bulk RNA-seq cohort (n=12) (Supplementary Table S3), a lower baseline activity score of the protein-translation program (GEP1) was similarly associated with suboptimal hematologic responses and was more active in patients without t(11;14) than in those with t(11;14) (Supplementary Fig. S4c). We also examined the treatment- and response-associated changes in GEP activity across the full cohort and within patient subgroups with and without t(11;14), revealing highly similar patterns across these analyses (Supplementary Fig. S4b).

The increase in mitotic signature in residual PCs led us to hypothesize that mitotic PCs may constitute a reservoir of proliferative, stem cell–like PCs that remains after treatment. To test this hypothesis, we tracked residual PC clones by linking pre- and post-treatment plasma cells based on their light-chain CDR3 sequences, which served as clone identifiers (Fig. 3d). In patient ALl5, who achieved a partial response (PR), five clones were detected at both baseline and after treatment, including one amyloidogenic PC clone. In patient ALk3, who achieved a VGPR, only one clone persisted after treatment, corresponding to the amyloidogenic PC clone. By contrast, patient ALl9, who achieved a complete hematologic response (CR), had no residual amyloidogenic PCs after treatment (Fig. 3e).

The clone tracking with light-chain CDR3 sequences allowed us to do longitudinal analysis of transcriptional state changes within the same clone across treatment. First, we found that PC clones from the same patient demonstrated highly similar copy number variation (CNV) patterns before and after treatment, per inferCNV results (Fig. 3f), indicating that AL may be genetically stable at the CNV level. We then performed DEG by comparing pre- and post-treatment cells with the same clone ID. In patient ALl5, the amyloidogenic clone exhibited a post-treatment transcriptional shift, characterized by increased stress adaptation and cytoskeletal remodeling programs and decreased protein translation activity. In patient ALk3, amyloidogenic plasma cells were nearly eliminated after treatment, with only 14 residual PCs detected. These residual cells showed increased inflammatory response programs and MHC class I gene expression, accompanied by reduced protein translation and cell-cell adhesion programs (Fig. 3g).

At the single-cell level, we classified PCs into three GEP-defined states: a normal PC state (GEP2^+^), a mitotic state (GEP5^+^), and a composite group comprising the other pathogenic states (Fig. 3h). This mitotic plasma cell state (GEP5^+^) was featured by high inferred stemness and G2M/S phase cell cycle scores (Supplementary Fig. S4d). In this representative patient with PR, clone tracking analysis indicated that PC clones were more likely to transition into mitotic states after treatment (Fig. 3i). Consistent with this finding, the proportion of cells expressing the mitotic GEP increased after treatment (Fig. 3j). Residual PCs in the mitotic state were predominantly localized to a small UMAP subpopulation characterized by high GEP5 activity and inferred stemness with CytoTRACE^17^ (Fig. 3j).

In summary, we identified PC GEPs whose activities were either associated with hematologic response or changed with Dara-Vd treatment, including a mitotic program enriched in residual PCs, in patients with t(11;14). Clonal dynamics analysis further supports a model in which mitotic PC clones act as a persisting reservoir that can give rise to other pathogenic PC states in patients with suboptimal hematologic responses.

### Prostaglandin E_2_-EP2/EP4 signaling and non-classical MHC class I signaling define an inflammatory–immunosuppressive bone marrow niche in suboptimal responders

In the following sections, we broadened our analysis to the BM immune microenvironment, focusing on immune cells interacting with PCs, to test the hypothesis that inflammatory–immunosuppressive dysregulation in the BM underpins suboptimal responses. We integrated several machine learning–based algorithms to dissect the BM microenvironment into 58 bone marrow cell types from all the whole BM samples (Fig. 4a). We manually curated B cells (Supplementary Fig. S5a, S5b), myeloid cells (Supplementary Fig. S5c), NK cells (Supplementary Fig. S5d), and T cell subtypes (Supplementary Fig. S5e).

**Fig. 4.**
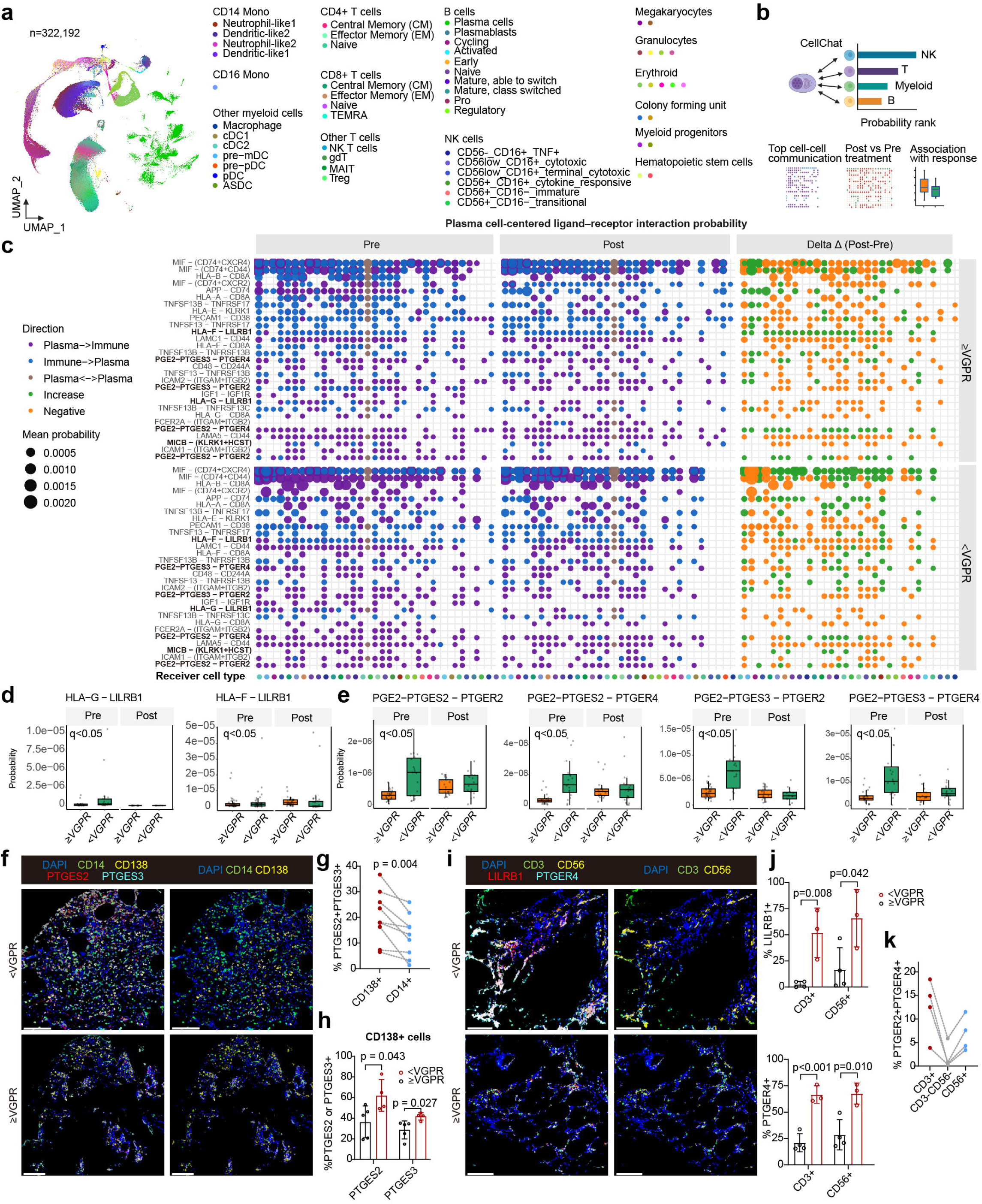
Plasma cell–centered immune cell communication in bone marrow niches. **(a)** Uniform manifold approximation and projection (UMAP) visualization of bone marrow (BM) plasma cells (PCs) and immune microenvironment cells in the whole BM samples. **(b)** PC–centered immune cell communication algorithm to compute the hematologic response-related ligand-receptor cell-cell communication probabilities. **(c)** Dot plot showing response-related ligand–receptor interactions between PCs and microenvironment immune cells grouped by pre- and post-treatment status and hematologic response. **(d)** Non-classical MHC class I signaling (*HLA-F–LILRB1* and *HLA-G–LILRB1* ligand– receptor pairs) associated with hematologic response. **(e)** Prostaglandin E_2_-EP2/EP4 signaling (*PTGES2/PTGES3*-*PTGER2/PTGER4* ligand-receptor pairs) associated with hematologic response. **(f)** Representative multiplex immunofluorescence images of BM sections from good and suboptimal responders, stained for DAPI, CD14, CD138, PTGES2, and PTGES3. Scale bars: 100 μm. **(g)** Percentages of PTGES2⁺PTGES3⁺ cells among CD138⁺ PCs and CD14⁺ myeloid cells in all BM samples (n = 8 per group). **(h)** Percentages of PTGES2⁺ or PTGES3⁺ CD138⁺ PCs in good and suboptimal responders (n = 5 and 4, respectively), quantified by multiplex immunofluorescence. **(i)** Representative multiplex immunofluorescence images of BM sections in good and suboptimal responders, stained for DAPI, CD3, CD56, LILRB1, and PTGER4. Scale bars: 100 μm. **(j)** Percentages of LILRB1⁺ and PTGER4⁺ cells among CD3⁺ T cells and CD56⁺ NK cells in good and suboptimal responders (n = 4 and 3, respectively), quantified by multiplex immunofluorescence. **(k)** Percentages of PTGER2⁺PTGER4⁺ cells across the indicated CD3⁺ and CD56⁺ T- and NK-cell populations in suboptimal responders (n = 4 per group).

To identify the immune processes most closely linked to treatment and response, we developed a PC-centered cell-cell communication analysis pipeline on whole BM samples. This analysis quantified ligand-receptor interactions between PCs and surrounding immune cell populations and then prioritized those interactions that changed most during treatment or differed by hematologic response category (Fig. 4b). Plasma cell interactions with myeloid, NK, T, and B cells collectively formed enhanced inflammatory and immunoregulatory niches associated with suboptimal hematologic responses, including *MIF–CD74/CD44/CXCR2 signaling*, adhesion via *ICAM1/2–ITGAM/ITGB2*, *LAMA5/LAMC1–CD44*, and *PECAM1–CD38*, non-classical MHC class I signaling with *LILRB1,* B-cell survival and maturation via *TNFSF13/TNFSF13B–TNFRSF13B/13C/17*, NK-cell/T-cell co-stimulatory signaling via *CD48–CD244A*, and prostaglandin (PG) signaling through PGE2–PTGER2/4 axes (Fig. 4c; Supplementary Table S4).

Across the pathways that were differentially active by hematologic response, non-classical MHC class I signaling and PG signaling were particularly enriched in suboptimal responders, both at baseline and after treatment. We observed an immunosuppression axis characterized by non-classical MHC class I cell-cell communication. PCs expressed *HLA-F* and *HLA-G*, which engaged inhibitory receptors such as *LILRB1* on NK cells, T cells, and myeloid cells (Fig. 4d). These non-classical MHC class I signaling pathways (*HLA-G*–*LILRB1* and *HLA-F*–*LILRB1*) were upregulated in patients with suboptimal responses at baseline, consistent with an immunosuppressive remodeling of the niche. In contrast, stress-induced MICB– (KLRK1+HCST) signaling between PCs and cytotoxic T cells was more strongly upregulated in good responders at baseline and was mechanistically associated with enhanced cytotoxicity mediated through the activating receptor KLRK1, consistent with effective antitumor immunity (Supplementary Fig. S6a).

In addition to immunosuppression by non-classical MHC class I pathways, PG-mediated inflammation was more active in patients with suboptimal hematologic responses than in those with good responses at baseline. Over the course of therapy, PG-mediated inflammation increased in patients with better responses and declined in those with suboptimal responses; however, PG-related inflammation remained higher in the suboptimal-response group, indicating a persistent PG-rich inflammatory niche in these patients, and that Dara-Vd may dampen but not reverse PG-mediated inflammation (Fig. 4e).

To further validate the PG-mediated cell–cell communication pathway, we performed multiplex immunofluorescence to resolve PG pathway components in BM. In the PTGES2/PTGES3/CD138/CD14 panel (Supplementary Fig. S7a-b), PTGES2 and PTGES3 were more frequently detected and co-localized in CD138⁺ PCs than in CD14⁺ monocytes (Fig. 4f-g; Supplementary Fig. S7c), indicating preferential enrichment of PG-synthesizing enzymes in PCs. Moreover, the proportions of PTGES2⁺ and PTGES3⁺ PCs were significantly higher at baseline in suboptimal responders than in good responders (Fig. 4f, 4h), supporting an association between enhanced PG pathway activity and suboptimal hematologic response.

Using a separate CD3/CD56/LILRB1/PTGER4 panel, we further characterized the T- and NK-cell populations receiving PG-mediated signals (Supplementary Fig. S8a-b). Consistent with the transcriptomic findings, the inhibitory receptor LILRB1 was preferentially enriched in CD56⁺ cells compared with CD3⁺ and CD3⁻CD56⁻ cells (Supplementary Fig. S8c). Moreover, both LILRB1 and PTGER4 were more frequently expressed in CD3⁺ T cells and CD56⁺ NK cells from suboptimal responders than in the corresponding populations from good responders (Fig. 4i-j). PTGER2 and PTGER4 co-localization was also confirmed in CD3⁺ T cells and CD56⁺ NK cells from suboptimal responders, suggesting that these receptors may have complementary roles in mediating PG signaling (Fig. 4k; Supplementary Fig. S8d).

Collectively, our analysis delineated a PC-centered cell-cell communication network in which the prostaglandin E_2_ (PGE₂)–EP2/EP4 (*PTGES2/PTGES3*-*PTGER2/PTGER4*) axis and non-classical MHC class I (*HLA-G*, *HLA-E*, and *HLA-F*) signaling were upregulated in patients with suboptimal hematologic responses at baseline, indicating a pre-existing inflammatory and immunosuppressive BM niche that may undermine the efficacy of Dara-Vd.

### Prostaglandin signaling was associated with negative regulation of interferon response in the bone marrow microenvironment

Previous studies have shown that chronic PG signaling can suppress IFN-γ responses across multiple immune-cell populations^18^. However, a key component of daratumumab-mediated anti–plasma-cell activity is the activation of cytotoxic lymphocytes that release IFN-γ^8^. We therefore sought to determine how PG signaling relates to the response to IFN-γ in AL during daratumumab-based treatment.

To further characterize the transcriptomic phenotype associated with PG signaling, we developed a multilayer graph network (ALNicheGraph). This graph contains three principal node types: cell nodes representing cell types, phenotype nodes representing ssGSEA functional enrichment terms, and signaling nodes representing CellChat ligand–receptor pairs. These nodes are connected by edges derived from pathway enrichment activity scores, ligand–receptor probabilities, or Spearman correlation coefficients (Fig. 5a; Supplementary Fig. S9a). When a signaling pathway is entered as a query, the direct-neighbourhood algorithm prioritizes phenotypes directly connected to the query, whereas personalized PageRank propagates the query through weighted paths in the graph to identify phenotypes connected through intermediate nodes (Fig. 5b).

**Fig. 5.**
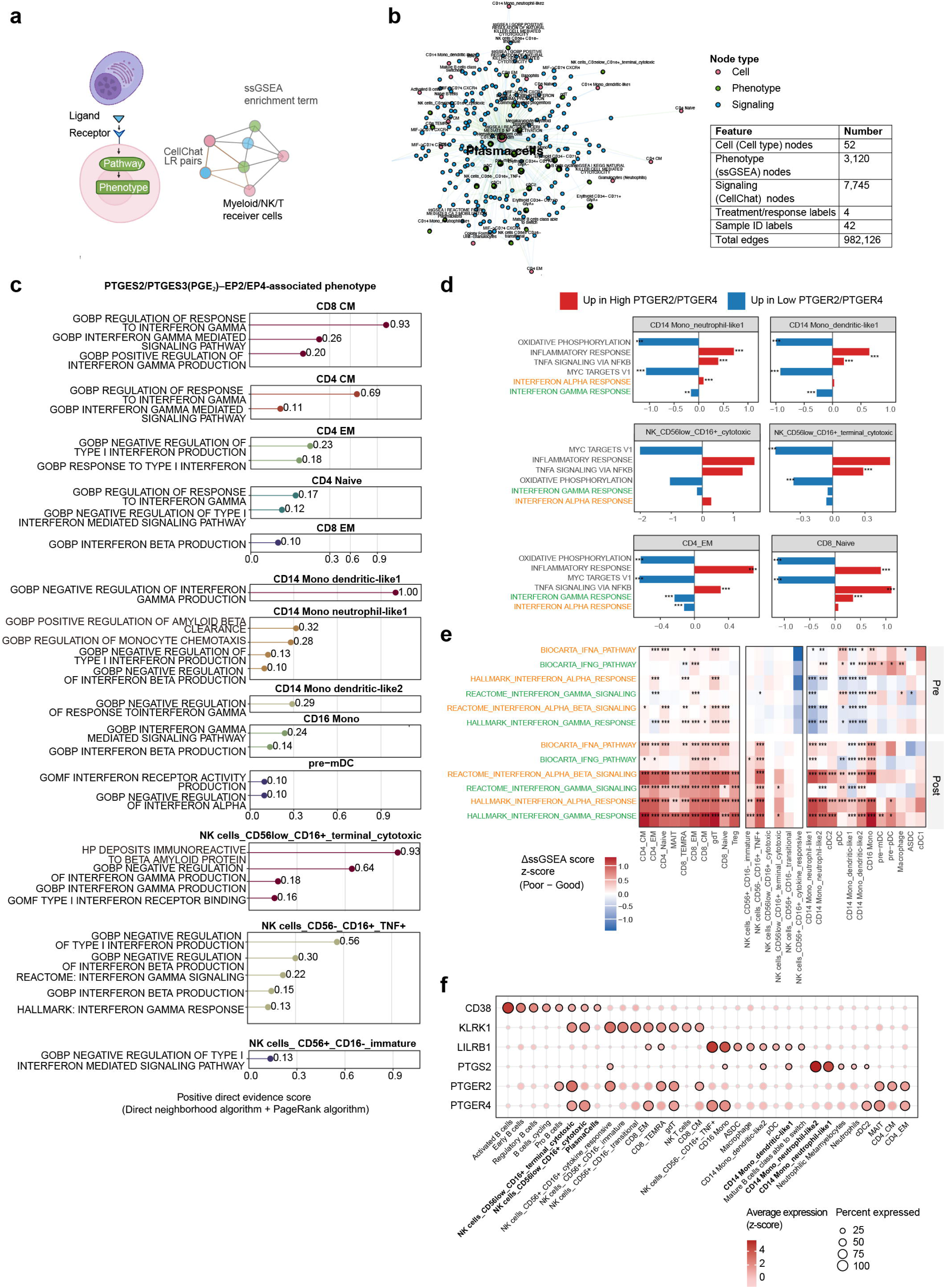
Prostaglandin signaling and its association with interferon responses in the microenvironmental immune cells. **(a)** Multilayer graph network to link cell-cell signaling to downstream pathway enrichment in microenvironment immune cells. **(b)** Network visualization of top-ranking signaling and phenotype axes in the graph-analysis framework. **(c)** Immune phenotypes positively linked to Prostaglandin E_2_–EP2/EP4 signaling identified by ALNicheGraph. **(d)** Heatmap showing differences in ssGSEA activity scores for interferon response between good and suboptimal responders, stratified by pre- and post-treatment conditions. **(e)** Mean log-normalized expression of CD38, non-classical MHC class I receptors, and prostaglandin pathway genes across BM microenvironment cells, highlighting top-expressing cell types.

ALNicheGraph enabled us to identify the top-ranked immune phenotypes associated with activation of the PG signaling pathway in AL. Consistently, increased PG signaling was usually associated with negative regulation of IFN-γ response in multiple subpopulations of T cells, myeloid cells, and NK cells. Notably, it was also positively linked to positive regulation of amyloid-β clearance and monocyte chemotaxis in CD14^+^ neutrophil-like monocytes and immunoreactivity to amyloid-β protein in terminal cytotoxic NK cells (Fig. 5c).

We also stratified the immune cells according to the expression level of *PTGER2/PTGER4*. Immune cells with detectable PG receptors were classified into high-, intermediate-, and low-expression tiers according to PG expression levels (Supplementary Fig. S10a)^19^. This allowed us to compare *PTGER2/PTGER4*^high^ cells with the *PTGER2/PTGER4*^low^ populations within each immune cell type. Consistent with previous studies, *PTGER2/PTGER4*^high^ T, NK, and myeloid cells exhibited increased inflammatory response and TNF-α–NF-κB signaling activities and reduced oxidative phosphorylation and MYC target V1 activity. Notably, IFN-γ response activity was lower in *PTGER2/PTGER4*^high^ cells than in *PTGER2/PTGER4*^low^ cells across most immune-cell subpopulations, whereas associations with IFN response activity were less consistent. These findings are consistent with the established inverse relationship between PG signaling and IFN-γ response (Fig. 5d; Supplementary Fig. S10b).

Despite cell type–specific differences at baseline, PG signaling and IFN-γ responses exhibited a consistent treatment-associated relationship. Compared with good responders, suboptimal responders, who had a stronger PG-associated inflammatory microenvironment (Fig. 4e), showed moderately higher IFN-γ response activity in T cells, comparable activity in NK cells, and lower activity across myeloid cells at baseline (Fig. 5e). Following treatment, however, IFN-response programs became substantially more activated in suboptimal responders than in good responders (Fig. 5e). This transition coincided with a marked reduction in PG signaling in suboptimal responders, whereas PG signaling increased slightly in good responders (Fig. 4e).

These findings support a coherent model: baseline elevated PG signaling reflects an immune response to amyloid deposition and is associated with suppression of IFN-γ response, particularly in suboptimal responders. During Dara-Vd treatment, PG signaling declined markedly in suboptimal responders, potentially relieving this inhibitory effect and permitting a stronger IFN-γ response than that observed in good responders. However, this enhanced IFN-γ activity did not translate into more effective anti–plasma-cell immunity.

### Mapping signaling senders and receivers in the bone marrow microenvironment provides insights for immune function analysis

Guided by these response-associated pathways, we examined the expression of signaling ligands, receptors, and upstream precursor synthesis involved in non-classical MHC class I and PG signaling across all cell types in the BM microenvironment. This analysis enabled us to identify the immune cell subpopulations that participate in the PG-mediated inflammation and non-classical MHC class I signaling in suboptimal responders.

CD38, the molecular target of daratumumab, was most highly expressed in B cell lineage, including PCs, as well as in cytotoxic NK cells, particularly the CD56^low^ CD16^+^ terminally differentiated subset (Fig. 5f). *CD38* expression was also detected in more than 25% of cells within each hematopoietic progenitor, dendritic-cell, and macrophage subpopulations (Supplementary Fig. S11a). This comprehensive survey guided our subsequent analyses of *CD38* expression changes in NK and myeloid cells and their associations with treatment response.

Prior knowledge indicates that PGE₂–EP2/EP4 signaling involves PGE₂ generated by PCs through PTGES2 and PTGES3 enzymes^20^, and sensed by the downstream myeloid cells, NK cells, and T cells through the receptors PTGER2 and PTGER4. In our data, *PTGS2* (*COX-2*), which converts PG precursors into PGH, was most abundantly expressed in CD14⁺ monocytes upstream of this PG axis (Fig. 5f). This implicates these populations as key sources of PG signaling that warrant further investigation. Expression of PGE₂ receptors, *PTGER2* and *PTGER4,* also showed cell-type specificity, thereby shaping which immune subsets are most likely to receive and execute the downstream effects of this axis. *PTGER2* was highly expressed in CD56^low^CD16^+^ cytotoxic/terminal cytotoxic NK cells, CD56^+^ CD16^+^ cytokine responsive NK cells, γδ T cells, CD8⁺ terminally differentiated effector memory (TEMRA) T cells, CD8⁺ central memory/effector memory cells, and CD4⁺ central memory/effector memory T cells, while *PTGER4* was enriched in the cell types overlapping with those highly expressing *PTGER2*, with preferential expression in CD56^low^CD16^+^ cytotoxic NK cells, CD56^-^ CD16^+^ *TNF*⁺ NK cells, CD16⁺ monocytes (Fig. 5f).

Similarly, receptors downstream of non-classical MHC class I signaling showed cell type–specific expression patterns: *KLRK1* was primarily expressed by CD56^low^ CD16⁺ NK cells and CD8⁺ T cell subtypes, whereas *LILRB1* was predominantly detected on CD8⁺ effector memory and TEMRA cells, cytokine-responsive NK cells, CD16⁺ monocytes, macrophages, and CD14⁺ dendritic-like monocytes (Fig. 5f).

Taken together, these receptor distributions provided a rationale to focus subsequent analyses on the effector and target cell subpopulations that mediate immune dysfunction in suboptimal responders.

### Myeloid-derived suppressor cell–like CD38**⁻** CD14**⁺** monocytes act upstream in a prostaglandin axis linked to reduced functions in myeloid cells

We then focused on myeloid cells, particularly CD14⁺ monocytes, which expressed *PTGS2*, the enzyme that converts PG precursors into PGH₂ upstream of the PGE – EP2/EP4 signaling axis. Our analysis encompassed the entire myeloid lineage, from hematopoietic stem and early progenitor cells through the monocyte and granulocyte branches to terminal dendritic cells and macrophages (Fig. 6a). CD14⁺ monocytes segregated into two major transcriptional states: neutrophil-like and dendritic-like^21^ (Fig. 6b; Supplementary Fig. S12a). The neutrophil-like CD14⁺ monocytes were CD38^-^ and exhibited features of monocytic MDSCs: *ITGAM*^+^ *CD15 (FUT4)*^−^ and *HLA-DR*^low^, and highly expressing *PTGS2*, providing a source of precursors for the PGE₂– EP2/EP4 axis (Fig. 6c).

**Fig. 6.**
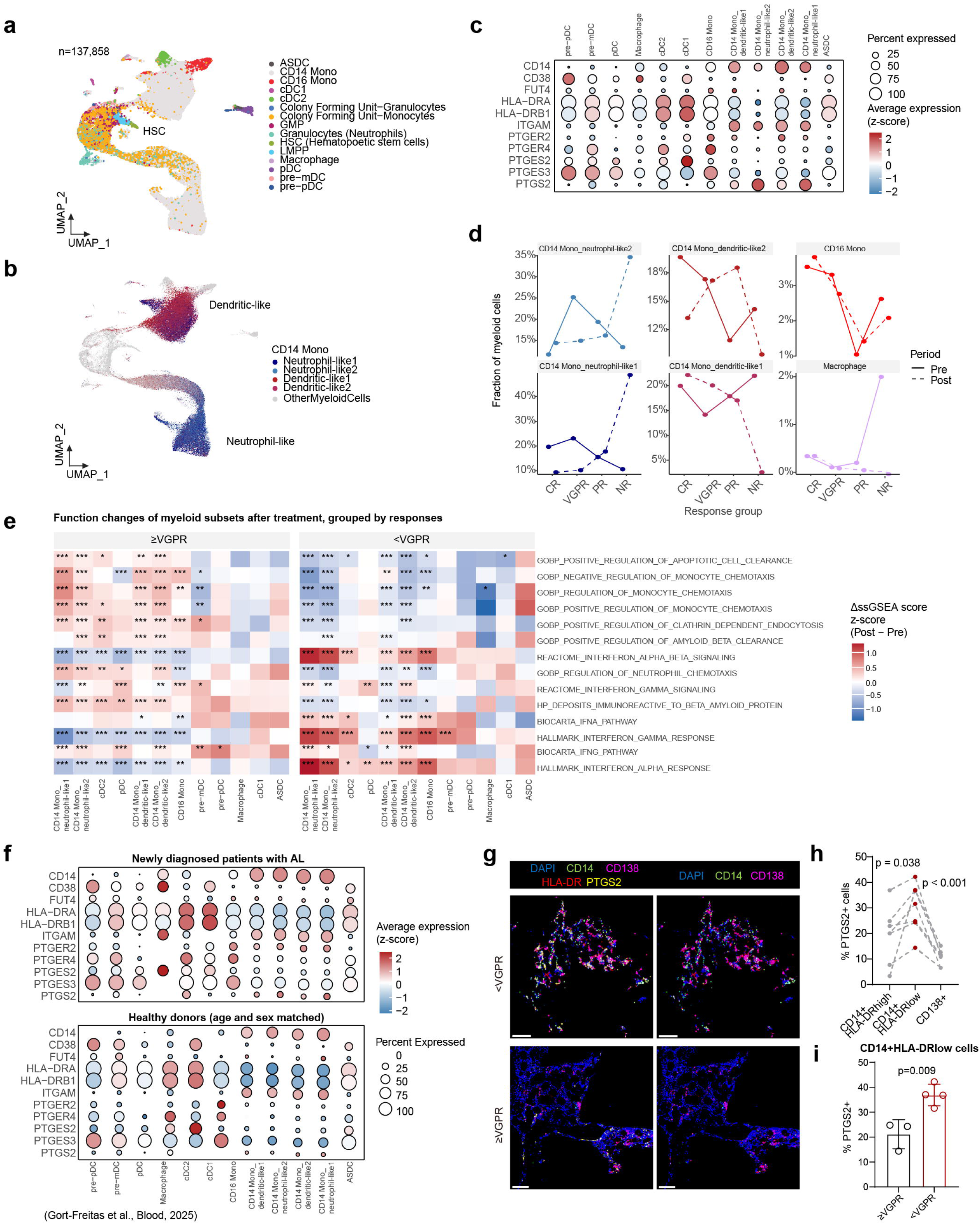
Myeloid-derived suppressor cell-like CD14**⁺** monocytes as upstream of prostaglandin signaling. **(a)** Uniform manifold approximation and projection (UMAP) visualization of myeloid lineage cells, from hematopoietic progenitors through mature states. **(b)** Four functional states of CD14 ⁺ monocytes, defined by the signatures of neutrophil chemotaxis/monocyte chemotaxis. **(c)** Mean log-normalized expression of myeloid-derived suppressor cell (MDSC) marker genes and prostaglandin pathway genes. **(d)** Changes in proportions of CD14 monocytes, CD16 monocytes, and macrophages following treatment, stratified by hematologic response. **(e)** Heatmap showing treatment-related changes in ssGSEA activity scores related to myeloid cell functions and interferon responses, stratified by hematologic response. **(f)** Mean log-normalized expression of MDSC marker genes and prostaglandin pathway genes in myeloid cells in patients with AL versus age- and sex-matched healthy donors (Gort-Freitas et al., Blood, 2025). **(g)** Representative multiplex immunofluorescence images of BM sections in good and suboptimal responders, stained for DAPI, CD14, CD138, HLA-DR, and PTGS2. Scale bars: 100 μm. **(h)** Percentages of PTGS2⁺ cells among CD14⁺HLA-DR^high^, CD14⁺HLA-DR^low^, and CD138⁺ cell populations in all BM samples (n = 6 per group). **(i)** Percentages of PTGS2⁺ cells in bone marrow samples from good and suboptimal responders (n = 3 and 4, respectively), quantified by multiplex immunofluorescence.

We next investigated how these *PTGS2*-expressing MDSC-like *CD38*⁻ CD14⁺ monocytes related to hematologic response to Dara-Vd. This subpopulation expanded markedly after treatment in suboptimal responders, although its baseline frequency did not significantly differ by response, a pattern that was paralleled by macrophages. By contrast, CD16⁺ monocytes were present at a significantly higher proportion at baseline in good responders than in suboptimal responders, and remained stable during treatment (Fig. 6d), suggesting that effective responses arise in a myeloid context enriched for CD16⁺ monocytes with surveillance functions. These findings show that the efficacy of Dara-Vd was associated with whether treatment is accompanied by a CD16⁺ monocyte–dominated, surveillance phenotype versus an inflammatory, MDSC-like CD14⁺ phenotype that fuels PG signaling. Additionally, despite being CD38⁺ and therefore susceptible to depletion by daratumumab (Fig. 6c), macrophages expanded markedly in suboptimal responders during treatment, even though their baseline proportions were comparable across response groups (Fig. 6d).

Within this inflammatory–immunosuppressive microenvironment, the myeloid effector functions that normally support ADCP were attenuated in patients with suboptimal responses. We observed that gene programs related to apoptotic cell clearance, clathrin-dependent endocytosis, amyloid-β clearance, and removal of amyloid-β– reactive deposits were downregulated, whereas IFN-γ response activity was upregulated. The most affected cell types were neutrophil-like CD14⁺ monocytes, CD16⁺ monocytes, and dendritic-like CD16⁺ monocytes, known to be the receiver cells of PG signaling and non-classical MHC class I signaling, as demonstrated by previous analysis. These results indicate that sustained exposure to PG and non-classical MHC class I stimulation may contribute to myeloid cell dysfunction (Fig. 6e).

Having identified the MDSC-like, PTGS2-expressing CD14⁺ neutrophil-like monocyte subpopulation, we next asked how specific it is to AL. In a single-cell dataset including patients with AL and age- and sex-matched controls (Supplementary Fig. S12b), the corresponding CD14⁺ neutrophil-like monocytes displayed a similar MDSC-like phenotype in patients, whereas the loss of *FUT4* expression was less consistent in healthy donors. In this monocyte subpopulation, *PTGS2* expression was also higher in AL samples than in matched healthy controls (Fig. 6f; Supplementary Fig. S12c)^9^. CD14⁺ neutrophil-like monocytes were likewise identified in a single-cell dataset of 98 healthy BM samples compiled from seven independent studies (Supplementary Fig. S12d)^22^. These cells again showed less consistent loss of *FUT4* expression (Supplementary Fig. S12e). The findings in external datasets indicate that CD14⁺ neutrophil-like monocytes are a normal component of the BM, whereas their MDSC-like features and elevated *PTGS2* expression are associated with the disease AL.

Using a panel comprising CD14/HLA-DR/PTGS2/CD138 (Supplementary Fig. S13a-b), we characterized this *PTGS2*⁺ MDSC-like CD14⁺ monocyte subset we observed in the transcriptome. PTGS2 signal was significantly enriched in CD14⁺HLA-DR^low^ cells compared with CD14⁺HLA-DR^high^ cells and CD138⁺ PCs (Fig. 6g-h), indicating that PTGS2 expression preferentially co-occurs with CD14 monocytes with an MDSC-like phenotype. Moreover, the proportion of PTGS2⁺ cells within the CD14⁺HLA-DR^low^ population was significantly higher at baseline in suboptimal responders than in good responders (Fig. 6g, 6i), supporting an association between an activated PG pathway and suboptimal hematologic response to a Dara-Vd.

To summarize, patients with suboptimal hematologic responses exhibited expansion of MDSC-like CD38^-^ CD14^+^ monocytes expressing PTGS2, which act upstream of the PGE –EP2/EP4 axis. Alongside this, the effector myeloid cells downregulated clearance of apoptotic cells and amyloid-β and upregulated response to IFN-γ, jointly establishing a dysfunctional inflammatory-immunosuppressive niche that undermines the efficacy of Dara-Vd.

### NK cells exhibit dysfunction and T cells show pronounced exhaustion, while both display increased IFN responses after treatment in suboptimal responders

Daratumumab exerts its immune effects through ADCC involving NK cells and ADCP mediated by macrophages/monocytes. Dysfunction of these pathways provides a conceptual basis for therapeutic resistance, particularly within chronically inflammatory and immunosuppressive microenvironments.

We next examined how Dara-Vd differentially shaped cytotoxic and exhaustion programs in NK cells and T cells between good and suboptimal responders, in the context of the inflammatory–immunosuppressive niches. Using functional markers and signatures, we identified six NK cell subtypes^23^ (Fig. 7a) and eleven T-cell subtypes^24^ (Fig. 7b). During Dara-Vd treatment, CD38⁺ NK cells and T cells (especially cytotoxic NK cells, γδ T cells, and CD8⁺ TEMRA cells) were depleted in all treatment response groups, whereas CD8⁺ effector-memory, CD4⁺ effector-memory, and CD4⁺ central-memory T cells expanded (Fig. 7c), indicating modulation of T and NK cell subpopulations by the regimen^6^.

**Fig. 7.**
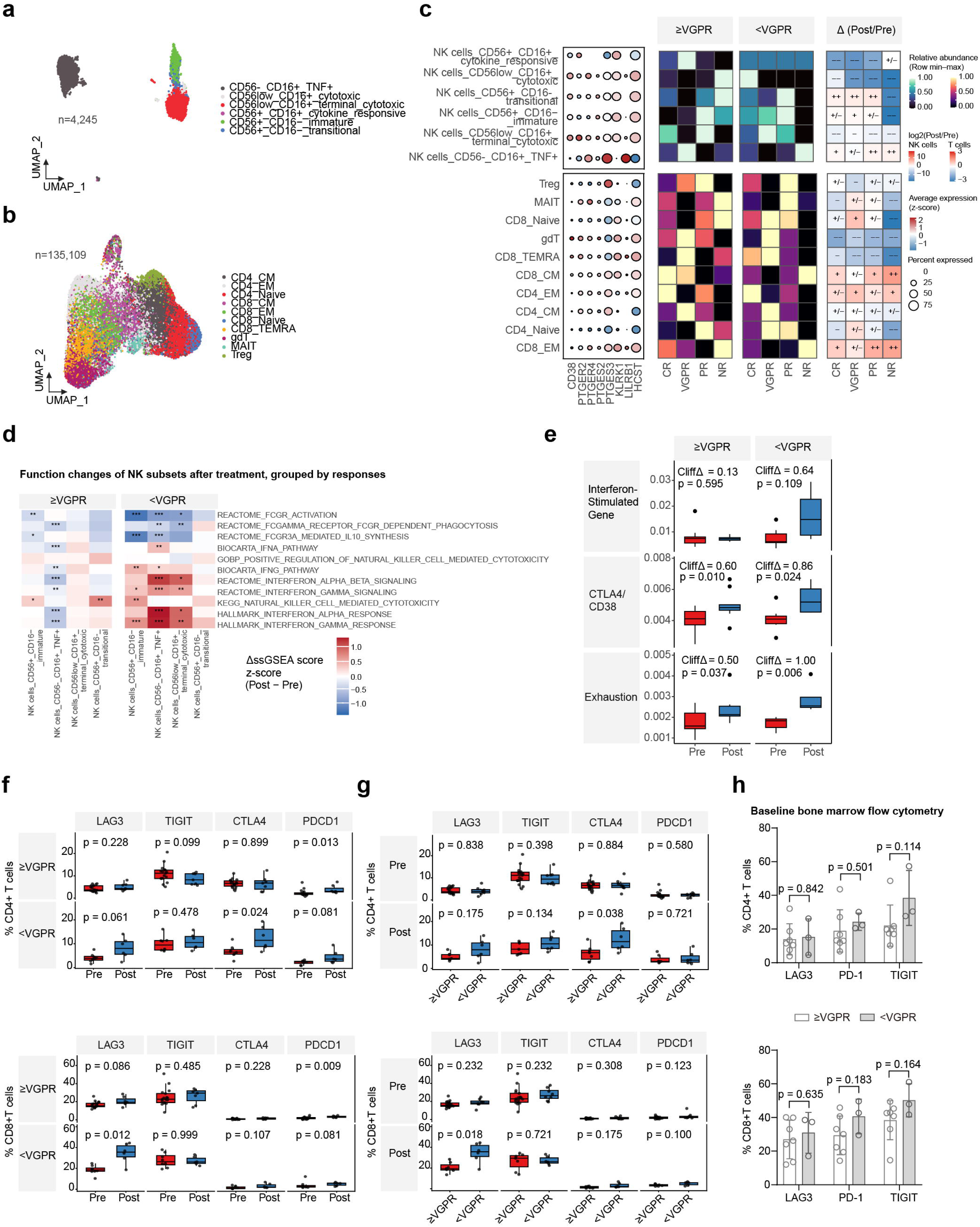
Increased interferon responses and exhaustion in NK-cell and T-cell lineages in suboptimal responders. **(a)** Uniform manifold approximation and projection (UMAP) visualization of NK cell subtypes in the BM microenvironment. **(b)** UMAP visualization of T cell subtypes in the BM microenvironment. **(c)** Mean log-normalized expression of genes in hematologic response–associated pathways and treatment-related changes in the proportions of T-cell and NK-cell subtypes, stratified by hematologic response. **(d)** Heatmap showing treatment-related changes in ssGSEA activity scores related to NK cell functions and inflammation, stratified by hematologic response. **(e)** Treatment-associated changes in T-cell functional signature scores grouped by responses. **(f)** Percentages of bone marrow CD4⁺ and CD8⁺ T cells positive for the exhaustion markers before and after treatment in patients with AL, stratified by hematologic response. **(g)** Percentages of bone marrow CD4⁺ and CD8⁺ T cells positive for the exhaustion markers in good and suboptimal responders before and after treatment. **(h)** Baseline percentages of CD4⁺ and CD8⁺ T cells positive for exhaustion markers (LAG3, TIGIT, and PD-1), measured by flow cytometry, stratified by treatment response (n = 7 and 3, respectively).

Within the immunosuppressive niche, NK cells exhibited transcriptional changes consistent with impaired ADCC induced by Dara-Vd. In suboptimal responders, CD56^low^CD16⁺ cytotoxic NK cells were selectively depleted; in good responders, this subset showed post-treatment upregulation of FcγR activation. Other NK cell subsets displayed a more pronounced downregulation of these effector programs in suboptimal responders compared with their counterparts in good responders. This pattern suggests a shift away from productive FcγR-dependent cytotoxic and regulatory functions toward a functionally dysregulated Fc-receptor state. Similar to myeloid cells, NK cells also exhibited increased IFN-γ response signatures (Fig. 7d), indicating that heightened IFN-γ signaling did not translate into anti–plasma-cell effector activity.

T cells mirrored these features; subsets in suboptimal responders displayed enhanced IFN-γ responses, together with increased activity of the IFN-stimulated gene signature, T-cell exhaustion signature, and inhibitory *CTLA4/CD38* signature (Fig. 7e). We next examined whether T-cell exhaustion differed according to hematologic response. Longitudinal comparisons of pre- and post-treatment samples within each response group showed that, among suboptimal responders, the *LAG3*⁺ and *CTLA4*⁺ fractions increased significantly in CD4⁺ T cells after treatment, whereas the *LAG3*⁺ fraction increased significantly in CD8⁺ T cells (Fig. 7f). Cross-sectional comparisons between response groups showed that the frequencies of cells positive for most exhaustion markers did not differ significantly between good and suboptimal responders at baseline or after treatment (Fig. 7g).

We further validated baseline T-cell exhaustion markers at the protein level by flow cytometry (Supplementary Fig. S14a). To validate the performance of the flow cytometry panel and contextualize the bone marrow findings, we first examined peripheral blood (PB) samples from nine healthy donors and compared paired PB and BM samples from patients with AL. LAG3 protein was low in both CD4⁺ and CD8⁺ T cells, whereas the median frequencies of PD-1 and TIGIT⁺ T cells in healthy-donor PB were consistent with previously reported ranges^25^ (Supplementary Fig. S14b). As expected, the frequencies of LAG3⁺ CD4⁺/CD8⁺ T cells and PD-1⁺ CD8⁺ T cells were higher in the PB of patients with AL than in healthy controls, similar to observations in other cancers ^26^ (Supplementary Fig. S14b) and were further increased in the BM compared with paired PB samples from the same patients^27^ (Supplementary Fig. S14c). We then compared the proportions of LAG3⁺, PD-1⁺, and TIGIT⁺ cells within the CD4⁺ and CD8⁺ T-cell subpopulations between good and suboptimal responders and found no statistically significant differences (Fig. 7h). These findings suggest that T-cell exhaustion is greater in the bone marrow than in PB in patients with AL. The more pronounced T-cell exhaustion in the BM of suboptimal responders compared with good responders emerged only after therapy and was not evident at baseline.

In summary, in patients with suboptimal responses, NK cells and T cells were selectively depleted by Dara-Vd and exhibited dysfunctional immune responses despite a heightened IFN-γ pathway, including impaired Fc receptor-mediated cytotoxic programs in NK cells and pronounced exhaustion in T cells, collectively contributing to reduced ADCC.

## DISCUSSION

In this study, we generated a single-cell atlas of BM PCs and the surrounding microenvironment from a prospective cohort of patients with AL treated with frontline Dara-Vd. This atlas revealed the features of PC clones and cell-cell communication between PCs and immune cells, especially those related to hematologic responses to the regimen. Our findings provide evidence for the hypothesis that suboptimal hematologic responses are associated with two major factors: intrinsic transcriptional programs of amyloidogenic PC clones and inflammatory-immunosuppressive dysregulation in the BM.

Amyloidogenic PCs have two major biological features that are well captured by our scV(D)J-seq/LC-MS and GEPs: clonal expansion and synthesis of amyloidogenic light chains. Within the t(11;14) subgroup, we observed that lower baseline protein translation and higher ER stress and UPR activity in PC clones predicted suboptimal hematologic response, suggesting that an adaptive ER stress response supports the survival of malignant PCs. This interpretation is consistent with a prior single-cell cohort study in MM showing that malignant PCs with elevated ER stress and UPR activation are associated with treatment resistance, particularly in relapsed/refractory

MM. Intriguingly, protein translation programs were relatively preserved during treatment in suboptimal responders compared with good responders. This program is intrinsically more active in amyloidogenic PCs with 1q21 gain and is mechanistically linked to light chain production. Together, these findings suggest that PC survival reflects a balance between light-chain production and ER stress and UPR programs.

We identified a mitotic PC state that shared clonotypes with other amyloidogenic PCs, increased in relative abundance following Dara-Vd treatment, and was capable of transitioning into other PC states. However, this PC state did not exhibit marker gene expression characteristic of the stem cell–like subpopulation previously described in MM, although the definition remains controversial. These observations motivate a key question for future studies: whether the proliferative, stem cell–like PC clones act as relapse-initiating clones in AL. Addressing this will require longitudinal sampling with clonal lineage tracing, along with functional validation to directly test their proliferation properties.

In our cohort, *HLA-G–LILRB1* and *HLA-F–LILRB1* interactions were associated with suboptimal response, whereas *MICB*–(*KLRK1*+*HCST*) interactions were linked to good response. These findings support a functional divergence among non-classical MHC-I signaling that depends on the receptor, in which inhibitory *LILRB1*-mediated interactions may contribute to immune suppression, while activating *NKG2D* signaling may preserve cytotoxicity. They extend a recent study by Kim et al., which showed that CD4⁺ cytotoxic T cells directly kill MM cells in an NKG2D-dependent manner and that the abundance of NKG2D⁺ CD4⁺ cytotoxic T cells correlates with improved survival^28^.

Moreover, our results are consistent with current knowledge of non-classical MHC class I signaling. HLA-G, through interaction with LILRB1, has been reported to inhibit cytotoxic T cells, induce T-cell anergy, and modulate myeloid cells. By contrast, MICB is expressed by many human cancers as a result of cellular stress and can tag cells for elimination by cytotoxic NK cells through activation via NKG2D^29^. If validated in independent cohorts, these findings support a clinical approach in which non-classical MHC class I genotype and expression are assessed in pre-treatment PCs from AL patients (for example, via PCR-based assays) to predict response to Dara-Vd.

Inflammation in cancer is a timely topic, and among inflammatory mediators, PGE₂ generated through the PG pathway is particularly important in driving tumor progression^30^. In our data, we observed broad PG signaling between PCs and NK cells, T cells, and myeloid cells. Although the association between PG signaling and drug resistance has not been reported in AL and is rarely described in MM, an experimental study by Mahaweni et al. used PGE₂ to mimic an inflammatory tumor microenvironment in vitro and showed that PGE₂ reduced NK cell cytotoxicity against MM target cells^31^. In solid tumors, PGE₂–EP2/EP4 signaling has been shown to impair the activity of CD8⁺ T cells and to induce PTGS2 expression in inflammatory myeloid cells and mature immunoregulatory dendritic cells^19^.

We identified a CD14^+^ monocytic MDSC phenotype expressing PTGS2, which is thought to arise from prolonged exposure to cytokines during chronic inflammation and cancer and is capable of differentiating into tumor-associated macrophages^32^, which intriguingly aligns with the parallel expansion of MDSCs and macrophages in suboptimal responders in our data. Krejcik et al. reported that CD38-expressing immunosuppressive MDSCs are sensitive to Dara-Vd treatment, whereas in our study, the MDSCs were *CD38*^-^, which may contribute to their expansion under daratumumab treatment^6^.

We found that key components of the PG pathway are distributed across different cell types: *PTGS2* is mainly expressed in monocytes, *PTGES2/PTGES3* are predominantly expressed in PCs, while the *PTGER2/PTGER4* are in multiple immune cell types. These findings suggest the presence of spatially organized inflammatory niches in which the cells involved in the PG pathway are spatial neighbours. Spatial profiling of the BM^33^ will therefore be essential to validate these immune niches inferred from scRNA-seq data. To further address this question, we have initiated a spatial transcriptomic study of BM biopsy samples from suboptimal and good responders within the same cohort and in independent validation cohorts.

Our observations of elevated IFN-γ responses and T-cell exhaustion within an inflammatory–immunosuppressive niche parallel findings from several prior single-cell studies in MM. In a recent analysis of MM resistance to heterogeneous daratumumab-based regimens, Wang et al. found that daratumumab initially activates cytotoxic immune cells and promotes IFN-γ-associated anti–plasma-cell activity, but with prolonged exposure, persistent T-cell activation sustains an IFN-γ-rich microenvironment, promotes T-cell exhaustion, and impairs effector-cell function^34^. At a larger scale, Pilcher et al. analyzed single-cell sequencing data from multicenter MM cohorts and showed that heightened inflammatory states— characterized by elevated IFN-γ activity and proinflammatory monocytes—were associated with poor clinical outcomes^35^. Earlier single-cell studies by Tirier et al. likewise reported increased IFN signaling and T-cell exhaustion in relapsed/refractory MM, although the cellular source of IFN was not specified^36^. Finally, Ziccheddu et al. demonstrated that in relapsed/refractory MM treated with daratumumab-lenalidomide-dexamethasone regimens, patients with progressive disease exhibited persistent T-cell exhaustion throughout therapy, even though IFN responses were not directly assessed^37^.

Our findings suggest that baseline inflammatory and immunosuppressive features in the bone marrow may support improved stratification of patients with AL receiving daratumumab-based therapy. Several biomarkers were elevated at both the RNA and protein levels in patients who subsequently experienced suboptimal responses and were detectable by immunofluorescence in pretreatment samples. These observations provide a foundation for developing clinically applicable assays, including bone marrow immunostaining to identify patients at increased risk of a suboptimal hematologic response. However, this strategy remains investigational, as no diagnostic assay has been validated in a larger cohort, and its sensitivity and specificity have yet to be established. Accordingly, patients identified as potential suboptimal responders at baseline could potentially benefit from adjunctive strategies targeting PG signaling. This possibility remains hypothesis-generating and requires rigorous mechanistic investigation and prospective clinical validation.

Our study has several limitations. First, single-cell profiling was restricted to freshly collected BM aspirates, precluding biobanking and introducing batch effects that are difficult to fully quantify. To mitigate this, we restricted differential analyses within samples and performed GEP analysis for PCs, which biologically have inter-patient heterogeneity that is hard to fully separate from the batch effect. Second, at post-treatment time points, limited BM material and low PC fractions often prevented adequate enrichment for 5′ scRNA-seq, resulting in relatively few post-treatment samples with matched CD138 ⁺ PC compartments. Third, although 1q21 gain has been implicated in suboptimal responses to daratumumab-based regimens by Chakraborty et al. (daratumumab-bortezomib-cyclophosphamide-dexamethasone)^4^ and Kimmich et al. (daratumumab-lenalidomide-dexamethasone)^38^, our limited cohort size left subgroup analyses underpowered, and we therefore did not observe a clear association in our data, despite an overall prevalence of 1q21 gain consistent with prior reports.

## CONCLUSIONS

In summary, this single-cell atlas identified two major bone marrow features associated with suboptimal hematologic responses to Dara-Vd: PC-intrinsic GEPs that support persistence of residual amyloidogenic clones after treatment, and baseline inflammatory–immunosuppressive niches characterized by PG and non-classical MHC class I signaling. These findings provide a resource and an analysis framework for identifying patients at risk for suboptimal response before treatment and for matching these patients to alternative therapeutic strategies based on the therapeutic targets present in their bone marrow microenvironment.

## METHODS

### Ethical approval

This study was approved by the *Ethics Review Committee of Peking Union Medical College Hospital, Chinese Academy of Medical Sciences*, and written informed consent was obtained from all participants (Approval No. JS-3455).

### Prospective and observational cohort

Patients were prospectively enrolled at Peking Union Medical College Hospital in a cohort with comprehensive baseline data and longitudinal follow-up, and samples were collected between March 2022 and June 2024. All enrolled cases met diagnostic criteria for AL as defined by current clinical guidelines and expert consensus^39,40^. Patients who had an involved-to-uninvolved free light chain ratio >100 at diagnosis and did not meet the diagnostic criteria of MM (Calcium elevation, Renal failure, Anemia, and Bone lesions, also known as CRAB features), were included based on expert adjudication supporting a primary diagnosis of AL. In addition, two healthy volunteers with normal BM were sampled as controls.

### Overview of bioinformatics analysis

A custom bioinformatics framework, PRISM (Plasma Cell Disease Repertoire Identification and Single-cell Mapping), was developed and utilized for the analysis. Briefly, we applied the CellRanger V(D)J pipeline and Dandelion^41^ to identify PC clonotypes and implemented an intra-patient differential gene–ranking algorithm to define GEPs of amyloidogenic PC clones. Subsequently, we employed cNMF^42^ and NNLS/starCAT^24^ algorithms to generate reference GEPs from clonal PCs and projected these programs onto all PCs, including those without captured clonotypes by single-cell V(DJ sequencing (scV(D)J-seq). For the paired pre- and post-treatment PC samples with scV(D)J-seq, we used the light-chain CDR3 sequence as the clone ID to match clones across time points, track clonal changes during treatment, and identify amyloidogenic PC clones. CytoTRACE^17^ was used to infer the CytoTRACE pseudotime, which represents the inferred stemness. InferCNVpy was used to infer clone-level copy number variation. We applied multiple machine-learning–based annotation tools, including SingleR^43^, NNLS/starCAT^24^, and Azimuth^44^, for annotating the BM microenvironment. Additionally, we formulated a PC-centered CellChat^45^ communication analysis to dissect the immune niches. Functional changes of individual immune cell types were evaluated using ssGSEA, thereby quantifying functional changes^46^ in immune cell states over the course of therapy.

### Detection of cytogenetic abnormalities

Isolated mononuclear cells were washed with hypotonic potassium chloride solution (0.075 M) and then Carnoy’s fixative (3:1 [v/v] methanol/acetic acid). The cell pellet was resuspended in Carnoy’s fixative and centrifuged again for additional washing. Interphase fluorescence in situ hybridization (FISH) was then performed on BM PCs using commercial probe kits (Abbott Molecular, Des Plaines, IL, USA). Positive thresholds for FISH results were defined according to the recommendations of the European Myeloma Network: t(11;14) > 10%, 1q21 gain > 20%, 17p (deletion) > 20%, and t(4;14) > 10%^47^. The hybridization signals were visualized under an OLYMPUS BX51 fluorescence microscope (OLYMPUS, Shinjuku City, Tokyo, Japan; Cat# WS-BX51-0169).

### 10X Genomics single-cell sequencing

We diluted 18 mL of BM aspirate with 18 mL of phosphate-buffered saline (PBS) and isolated mononuclear cells using Lymphoprep™ density gradient medium (STEMCELL Technologies, Vancouver, Canada; Cat# 18061) according to the manufacturer’s instructions. Each BM sample was split into two equal portions: one portion was sequenced for whole BM cells, and the other portion was enriched in PCs. PC enrichment was performed using CD138 magnetic beads (Miltenyi Biotec, Bergisch Gladbach, Germany; Cat# 130-051-301), MS columns (Cat# 130-042-201), and a MiniMACS separator (Cat# 130-042-102). The resulting PC suspensions were assessed for viability and quality using a LUNA-FL™ automated fluorescence cell counter (Logos Biosystems, Anyang-si, Gyeonggi-do, South Korea; Cat# L20001). Samples meeting the following criteria were processed for single-cell library preparation: cell viability > 85%, cell concentration 700–1200 cells/μL, diameter 5–10 μm, and total cell count > 1 × 10. We also ruled out cell aggregation, background impurities, and debris with the cell counter. 10X Genomics 3′ single-cell RNA-seq was performed on whole BM samples, while 5′ single-cell RNA-seq with paired scV(D)J-seq was performed on samples enriched with CD138 magnetic beads, following the standard library construction protocols. scV(D)J-seq captures the BCR sequence from RNA transcripts, which includes the light-chain sequences of amyloidogenic PCs. Loaded cell numbers were approximately 13,000 live cells for 3′ libraries and 12,000 for 5′ libraries. Library sequencing was carried out on Illumina NovaSeq X Plus (2023–2024) (Illumina, San Diego, CA; Cat# 20084804) and NovaSeq 6000 platforms (2022) (Illumina, San Diego, CA; Cat# 20012850) to a sequencing depth sufficient to reach saturation.

### Quality control and data pre-processing

Cell Ranger (version 6.0.1) was used to align FASTQ files. Single-cell gene expression matrices were generated using the Cell Ranger count pipeline from scRNA-seq data (reference: refdata-gex-GRCh38-2020-A), and the V(D)J sequence library was obtained using the Cell Ranger vdj pipeline (reference: refdata-cellranger-vdj-GRCh38-alts-ensembl-5.0.0). Seurat (version 5.1.0) was used to process the single-cell data matrix. A total of 78 scRNA-seq libraries were integrated using Seurat. Cells were retained if they contained more than 1,000 and fewer than 50,000 unique molecular identifiers (UMIs) and had mitochondrial transcript fractions below 25%. For each sample, the scRNA-seq gene expression matrix was log-normalised and then processed with variable selection, principal component analysis, k-nearest neighbors and the Louvain algorithm to identify cell clusters. DecontX^48^ was used to remove ambient RNA, and scDblFinder^49^ was applied to identify and exclude 62,864 doublets, resulting in a total of 630,111 high-quality single cells. Immunoglobulin transcripts excluded from the selection of highly variable genes and principal components for the embedding and signature selection, while being kept in the gene expression matrices for downstream analyses.

### Identification of plasma cell clonotype

Clonotypes of PCs were defined by Cell Ranger vdj based on identical full-length productive CDR3 nucleotide sequences of paired heavy (IGH) and light chains (IGL/IGK)^50^, respectively, allowing us to identify amyloidogenic PC clones according to the light chain restriction. CDR3 nucleotide identity ≥ 85% was required for clonotype determination. scV(D)J-seq libraries were then processed with Dandelion to standardize V/J calls, improve contig labels, and assemble full-length light chain sequences. The consensus full-length light-chain sequence was calculated by a custom pipeline from each PC clone, defined as the predicted amino acid sequence derived from the RNA transcript spanning the FWR1, CDR1, FWR2, CDR2, FWR3, CDR3, and FWR4 regions. For each patient and time point, we identified the dominant expanded PC clones by intersecting the scRNA-seq library with the V(D)J library. For these dominant clones, we summarized isotype usage (κ vs λ) and computed a consensus light chain sequence. PCs without a corresponding V(D)J clonotype assignment were considered technical dropouts and were not assigned a clonotype.

### Intra-patient differential gene ranking algorithm

Based on the clonotype and light chain sequence, PCs were categorized into amyloidogenic monoclonal PCs and normal polyclonal PCs. For each patient, a paired differential expression analysis was performed to compare amyloid clones versus the intrinsic normal control, yielding a list of differentially expressed genes (DEGs) specific to each patient. The DEG lists from all patients were then aggregated, and DEGs were ranked according to either their recurrence frequency across patients or their mean expression fold change, resulting in a prioritized list of high-frequency DEGs. Finally, the top 100 downregulated/upregulated genes were used for functional enrichment analysis and marker selection.

### Gene expression program analysis with cNMF and NNLS/starCAT

Consensus non-negative matrix factorization (cNMF) was applied to identify GEPs across single cells^42^. When given an scRNA-seq expression matrix (N cells × G genes), cNMF decomposes the data into two matrices: a (K × G) matrix representing the gene composition of each expression program, and an (N × K) matrix representing the activity of each program in individual cells. We built the GEP reference from clonally defined PCs, which ensured that the learned GEPs specifically reflected features of amyloidogenic versus normal PCs. GEPs that capture contamination signatures were removed. We subsequently applied non-negative least squares (NNLS) via the starCAT pipeline^24^ to map these reference programs onto all PCs, including the minimal residual disease PCs. This projection quantified the activity of GEPs across the entire PC population, enabling comparison of GEP activity between amyloidogenic PCs, polyclonal PCs, and minimal residual disease after treatment.

### Clonal copy number variation inference with InferCNVpy

InferCNVpy was used to estimate copy number variation (CNV) based on a single-cell data matrix. Polyclonal normal PCs served as reference control. Genes were mapped and ordered according to chromosome coordinates, and chromosomal CNV signals were calculated from the mean gene expression in a sliding window of 100 genes, with a minimum expression cutoff of 0.1 to filter lowly expressed genes before CNV estimation. For better visualization, we clustered CNV profiles by the p and q arms of chromosomes.

### Immune cell annotation with multiple machine learning algorithms

To systematically annotate immune and hematopoietic cell populations in the BM, we employed a multi-step annotation framework integrating several machine learning– based algorithms and manual curation. We first applied SingleR^43^ using the Novershtern Hematopoietic Data as the training reference to annotate all immune and hematopoietic cells except PCs. This initial classification identified 38 distinct cell types across the BM samples. Because the Novershtern dataset is derived from healthy volunteers, it may not fully capture disease-specific or fine-grained immune cell subtypes. To refine these annotations, we further analyzed specific immune lineages using specialized algorithms. For T cells, we applied starCAT with default parameters, which provides a signature-based classification of T cell functional states^24^. For myeloid cells, we utilized Azimuth^51^, an annotation algorithm based on the Human BioMolecular Atlas, to project our data onto a myeloid differentiation trajectory spanning hematopoietic progenitors to mature myeloid cells. Finally, B cells and NK cells were manually curated based on canonical lineage and functional markers to preserve disease-relevant phenotypes. Harmony was used to integrate scRNA-seq data from different patients within each immune cell type^52^.

### Plasma cell-centered cell-cell communication analysis with CellChat

To characterize the cellular interactions between PCs and the surrounding immune microenvironment, we performed a systematic ligand–receptor-level cell-cell communication analysis using CellChat^45^. We first isolated PCs and immune cell subsets from the whole BM samples, as these samples most accurately reflect the cell type composition of the BM niche. We briefly examined the expression of canonical marker genes, including CD38, to identify cell populations with high expression of the therapeutic target. Subsequently, CellChat was independently applied to each sample to infer ligand–receptor interactions. From these results, we extracted interaction pairs in which PCs served as either senders or receivers. Interaction probabilities were ranked after correction for the proportion of each immune cell subset. We then compared the PC–involving interactions between pre-treatment and post-treatment samples to delineate how communication patterns change over therapy, particularly in relation to hematologic response.

### ssGSEA functional enrichment for immune cell phenotypes

Single-sample Gene Set Enrichment Analysis (ssGSEA)^53^ is a modified version of the traditional GSEA algorithm that quantifies pathway activity at the single-cell level. ssGSEA computes an activity score for each gene set in each cell based on the ranked expression profile. The gene sets for enrichment were collected from the Molecular Signatures Database (MSigDB), focusing on pathways characteristic of each lineage including phagocytosis and endocytosis in myeloid cells, ADCC in NK cells, and major immune-response pathways in myeloid, T, and NK cell subtypes (C2:CP: BIOCARTA; C2:CP: REACTOME; C5:GO: BP; H: HALLMARK). The Wilcoxon test was used to test the significance of the comparison, and the change in activity scores, ΔssGSEA score (mean(Post) − mean(Pre)), was plotted.

### GSVA for calculating gene expression program scores from bulk RNA-seq samples

We performed whole transcriptomic sequencing on CD138⁺ BM cells enriched by CD138⁺ magnetic bead selection. The mRNA libraries were prepared with the VAHTS Universal V6 RNA-seq Library Prep Kit for Illumina (Vazyme biotech, Nanjing, China, Cat# NR604) and then sequenced on Illumina NovaSeq 6000 (Illumina, San Diego, CA; Cat# 20012850) if the RNA Integrity Number (RIN) was higher than 7. We applied Gene Set Variation Analysis (GSVA) to compute enrichment scores for each GEP in the samples from the bulk RNA-seq cohort (Supplementary Table S3). GSVA is a non-parametric, unsupervised method that estimates the relative enrichment of predefined gene sets across samples within an expression dataset^53^.

### ALNicheGraph for signaling-phenotype association analysis

We developed ALNicheGraph, a multilayer graph network integrating cell–cell communication, intracellular pathway activity, and functional phenotypes. The graph contained three principal node types: cell nodes representing cell populations, signaling nodes representing CellChat^45^ ligand–receptor interactions, and phenotype nodes representing functional enrichment terms derived from ssGSEA^46^. CellChat outputs were converted into directed signaling relationships specifying the sender cell type, receiver cell type, signaling pathway, ligand–receptor pair, and interaction probability. In parallel, ssGSEA pathway activity scores were calculated for each cell population and sample and used to connect cell populations with their corresponding functional phenotypes. Signaling–phenotype relationships were further quantified using Spearman correlations between signaling activity and phenotype enrichment, together with evidence from pre-versus post-treatment changes, and response-group differences. Because these data were generated at different levels of resolution, they were aligned using shared sample, cell-type, and signaling identifiers and integrated into the multilayer graph, with edge weights derived from CellChat interaction probabilities, pathway enrichment activity scores, or Spearman correlation coefficients. Clinical information was retained as sample context for context-specific graph queries.

For receiver-phenotype analyses, ALNicheGraph was queried according to the signaling pathway, receiver cell type, sample context, and treatment or response status. Phenotypes associated with a queried signaling pathway were ranked using either direct-neighbourhood analysis, which prioritizes phenotypes directly connected to the query, or personalized PageRank, which propagates the query through weighted paths in the graph to identify phenotypes connected through intermediate nodes. A combined ranking score integrating these approaches was additionally used where indicated. Query results were reported as ranked phenotype tables and corresponding local subgraphs.

### Validation experiment 1: Full-length LC–MS analysis of light-chain protein in serum samples

Immunoglobulin G was depleted from patients’ serum using Protein A/G Magnetic Beads (Abcam, Waltham, MA; Cat# ab286842) to enrich free light chains, including monomeric and dimeric forms. κ or λ FLCs were separately captured using CaptureSelect™ KappaXL Affinity Matrix (Thermo Fisher Scientific, Waltham, MA; Cat# 1943210250) and CaptureSelect™ LambdaXP Affinity Matrix (Thermo Fisher Scientific, Waltham, MA; Cat# 1943752250), respectively. Enriched fractions were separated by non-reducing SDS–PAGE (Bio-Rad Laboratories, Hercules, CA), and bands corresponding to FLC monomers and dimers were excised for in-gel digestion with Mass Spectrometry Grade Proteases, including Trypsin, Chymotrypsin, and LysC (Thermo Fisher Scientific, Waltham, MA; Cat# 90059 for Trypsin, #90056 for Chymotrypsin, and #90307 for LysC) to maximize sequence coverage. Digested peptides (0.5 μg) were loaded onto EV-2001 C18 Evotips (Evosep, Odense, Denmark; Cat# EV-2001) and separated on a PepSep™ C18 analytical column (length: 15 cm; inner diameter: 100 μm; particle size: 3 μm) (Bruker Daltonics, Billerica, MA; Cat# 1895806) using the 30 Samples Per Day (SPD) Evosep method. Mass spectrometry was performed on an Orbitrap Exploris Mass Spectrometer (Thermo Fisher Scientific, Waltham, MA; Cat# 1943210250) under previously described parameters.^54^ Raw data were analyzed using the REmAb™ De Novo protein sequencing platform (Rapid Novor Inc., Kitchener, ON, Canada) to de novo assemble full-length free light chain sequences for each patient.

### Validation experiment 2: Multiplex immunofluorescence staining of bone marrow samples

Patients with AL newly diagnosed at Peking Union Medical College Hospital were retrospectively retrieved from the pathology department. To investigate the distinct spatial phenotypes within the tissue microenvironment, formalin-fixed paraffin-embedded (FFPE) tissue sections were subjected to multiplex immunofluorescence staining using four distinct biomarker panels. The panels were designed as follows: Panel 1 (CD14, CD138, HLA-DR and PTGS2); Panel 2 (CD3, CD56, LILRB1 and PTGER4); Panel 3 (CD14, CD138, PTGES2 and PTGES3), and Panel 4 (CD3, CD56, PTGER2 and PTGER4) (Supplementary Table S5).

Initially, sections were deparaffinized in xylene (three times, 5 min each) and sequentially rehydrated through a graded ethanol series (100% twice, 95%, 90%, 80%, 70%, and 50% ethanol for 5 min each), followed by washing in distilled water. A hydrophobic barrier was drawn around the tissue using a PAP pen, and the slides were washed with PBS three times (5 min each). For the detection of the four target proteins within each panel, iterative cycles of staining were performed. Each cycle consisted of the following sequential steps: (1) Antigen retrieval and antibody stripping: Sections were treated with EDTA buffer (Solarbio, Beijing, China; Cat# C1034) in a pressure cooker for 5 min and allowed to cool to room temperature (RT). (2) Blocking: Endogenous peroxidase activity was quenched using a peroxidase blocking solution (Solarbio, Beijing, China; Cat# G2850) for 15 min at RT. After washing with PBS (three times, 5 min each), tissues were blocked with 3% bovine serum albumin (BSA) (Solarbio, Beijing, China; Cat# SL038) for 30 min at RT. (3) Primary antibody incubation: Sections were incubated with the specific primary antibody corresponding to the designated target in the panel cycle, in a humidified chamber at 4 °C overnight. The following day, slides were equilibrated to RT for 30 min. (4) Secondary antibody incubation: Slides were washed once with PBST (PBS containing 0.1% Tween-20) for 5 min and three times with PBS for 5 min each. Subsequently, sections were incubated with the corresponding species-specific secondary antibody for 1 h at RT in the dark, followed by three washes in PBS. (5) Signal amplification: Tyramide signal amplification (TSA) was performed by incubating the sections with the secondary antibody for 25 min at RT in the dark. Following TSA incubation, slides were washed once with PBST for 5 min and three times with PBS for 5 min each (Supplementary Table S6). This five-step cycle was repeated four times in total for each slide to sequentially label all four targets of the respective panel.

After completing the final cycle of TSA staining, cell nuclei were counterstained with DAPI (Invitrogen, Carlsbad, CA; Cat# D1306) for 15 min at RT in the dark. Finally, the slides were washed three times in PBS (5 min each) and coverslipped using an anti-fluorescence quenching mounting medium (Servicebio, Wuhan, China; Cat# G1401). Images were acquired using a Olympus VS200 whole-slide scanner (OLYMPUS, Shinjuku City, Tokyo, Japan; Cat# vs200).

Following image acquisition, the multi-channel digitized images were imported into QuPath software (version 0.7.0) for quantitative single-cell analysis. Cell segmentation was performed based on the DAPI nuclear counterstain using the built-in cell detection algorithm (setting sigma = 1 μm), followed by a 3-μm cellular expansion to approximate the cytoplasmic boundaries for each individual cell. Subsequently, the mean fluorescence intensities for all targeted markers within the defined cellular compartments were computed and extracted. Empirical thresholds for marker positivity were carefully established by evaluating the visual fluorescence background directly on the tissue images using the ‘Single measurement classifier’ tool in QuPath software. The quantified single-cell intensity data were subsequently imported into FlowJo software (version 10.8.1) (BD Biosciences, Woburn, MA; Cat#10.8.1). Within FlowJo, hierarchical gating strategies guided by the visually validated thresholds determined in QuPath were applied to identify specific cellular populations.

### Validation experiment 3: Flow cytometry analysis of T-cell exhaustion markers from peripheral blood and bone marrow samples

Patients with newly diagnosed AL at Peking Union Medical College Hospital and healthy individuals from a routine health screening cohort were enrolled during the same period (Supplementary Table S7).

PB and BM samples used for flow cytometry were collected before treatment initiation from all participants. For each participant, 2 mL of PB and/or BM was collected into EDTA tubes (BD Biosciences, Woburn, MA; Cat# 367841). For surface staining, 100 μL of sample was added to pre-labeled flow cytometry tubes containing 5 μL each of the following antibodies or reagents: CD4; PE anti-human (BioLegend, San Diego, CA; Cat#344606); PE anti-human CD197/CCR7 (BioLegend, San Diego, CA; Cat# 353204), PerCP anti-human CD3 (BioLegend, San Diego, CA; Cat# 317338), PE/Cyanine7 anti-human CD45RA (BioLegend, San Diego, CA; Cat# 304126), APC anti-human CD279/PD-1 (BioLegend, San Diego, CA; Cat# 329908), Alexa Fluor 700 anti-human CD223/LAG-3 (BioLegend, San Diego, CA; Cat# 369344), APC/Cyanine7 anti-human CD28 (BioLegend, San Diego, CA; Cat# 302966), Brilliant Violet 510 anti-human CD45 (BioLegend, San Diego, CA; Cat# 368526), Brilliant Violet 605 anti-human CD8 (BioLegend, San Diego, CA; Cat# 344742), PE/Dazzle 594 anti-human CD56/NCAM (BioLegend, San Diego, CA; Cat# 362544), Zombie Violet Fixable Viability Kit (BioLegend, San Diego, CA; Cat# 423114), and BV650 mouse anti-human TIGIT (BD Biosciences, Woburn, MA; Cat# 747840). Samples were mixed and incubated for 15 min at room temperature in the dark.

A permeabilization kit containing solutions A and B was used (Beijing Lifang Scientific, Beijing, China; Cat#26600808). After surface staining, 100 μL of permeabilization solution A was added to each tube, and samples were mixed and incubated for 15 min in the dark. Samples were then centrifuged at 1500 rpm for 5 min, washed once with 2 mL PBS, centrifuged again at 1500 rpm for 5 min, and resuspended by vortexing (Beckman Coulter, Brea, CA; Cat#75009510). After removal of the supernatant, 100 μL of permeabilization solution B was added, followed by 5 μL of intracellular anti-human CD152/CTLA-4-FITC antibody (Invitrogen, Carlsbad, CA; Cat# 11-1529-42). Samples were stained for 15 min in the dark, washed with 2 mL PBS, centrifuged at 1500 rpm for 5 min, and resuspended in 200 μL PBS. Flow cytometry was performed on a DxFLEX B5-R3-V5 Flow Cytometer (Beckman Coulter, Brea, CA; Cat# C44327 B5-R3-V5).

Flow cytometry data were analyzed using FlowJo software (version 10.8.1) (BD Biosciences, Woburn, MA; Cat#10.8.1). CD4⁺ and CD8⁺ T cells were defined as CD45^+^CD3^+^CD8^-^ and CD45^+^CD3^+^CD8^+^, respectively. Within each T-cell subset, the percentages of cells expressing PD-1, TIGIT, and LAG-3 were quantified.

## Statistical analysis and visualization

R (version 4.3.3) and Python (version 3.8.19) were used for statistical analyses and data visualization. In bioinformatics analyses, continuous variables were compared between two independent groups using the Wilcoxon rank-sum test and among more than two independent groups using the Kruskal–Wallis test, as appropriate. Categorical or count data were compared using the chi-squared test, where applicable. For analyses involving multiple hypothesis testing, p values were adjusted using the multiple-testing correction specified for the corresponding analysis, including the Benjamini–Hochberg false discovery rate (FDR) procedure. Statistical significance was defined as p < 0.05 for unadjusted analyses or FDR-adjusted p < 0.05 (q < 0.05) for analyses with multiple-testing correction. Unless otherwise specified, all statistical tests were two-sided. For flow cytometry and multiplex immunofluorescence analyses, comparisons between two groups were performed using paired or unpaired tests according to the study design. Paired t-tests were used for paired samples and unpaired t-tests for independent samples when parametric assumptions were appropriate. Otherwise, the corresponding nonparametric tests were used, including the Wilcoxon signed-rank test for paired samples and the Wilcoxon rank-sum test for independent samples. For comparisons involving more than two independent groups, the Kruskal– Wallis test was used.

## Code availability

Codes wrapped in Jupyter notebooks are available in the GitHub repository (https://github.com/Selecton98/SingleCell_PRISM_pALDaratumumab).

## Data availability

The raw sequencing data have been deposited in the Genome Sequence Archive for Human at the NGDC, CNCB (GSA-Human: single-cell data HRA017125, bulk RNA-seq HRA017315), and will be publicly accessible at https://ngdc.cncb.ac.cn/gsa-human upon publication. The processed single-cell object with annotation and sample information will be on Zenodo (https://zenodo.org/) upon publication. The de-identified clinical information for the patients is provided in Supplementary Table S1 (single-cell cohort), S3 (bulk RNA-seq cohort), S6 (multiplex immunofluorescence cohort) and S7 (flow cytometry cohort). The other data are available upon reasonable request to the corresponding author. Source Data are provided with this paper.

## Acknowledgements

X.W. thanks Dr. Fangyuan Chen (School of Medicine, Tsinghua University) for her suggestions on the results. The authors extend their sincere gratitude to the patients who participated in this study and their families. This study was funded by the National Natural Science Foundation of China (Grant No. 82470202 for LJ), the CAMS Innovation Fund for Medical Sciences (CIFMS) (Grant No. 2025-I2M-XHXX-010 for SKN), the National High Level Hospital Clinical Research Funding (Grant No. 2025-PUMCH-A-182 for SKN), and the Research Initiative for Post–Marketing Clinical Studies of Innovative Pharmaceuticals (Grant No. WKZX2024CX105204 for LJ).

## Author contributions

X.W. conceptualized the study, designed the methodology, analysed the data, and drafted the manuscript; X.X. analysed the data and revised the manuscript; C.X. performed the validation experiment, analysed the data, and revised the manuscript; H.H. collected the samples; A.G. and Y.G. performed the validation experiment; A.G., Y.G., and K.S managed the clinical cohort; Y.Q. revised the methodology and the manuscript; K.S. and J.L. conceptualized the study, supervised the project, and revised the manuscript. All the authors read and approved the final manuscript.

## Use of Artificial Intelligence in manuscript preparation

We enhanced the readability and clarity of this manuscript with assistance from a large language model artificial intelligence system, which was used to check grammar and spelling, improve sentence structure, and overall readability of the text while preserving the original scientific content and conclusions. Suggested revisions were reviewed and improved by the authors before incorporation into the manuscript. The use of AI assistance was limited to editorial improvements and did not involve the generation of research data, statistical analyses, or scientific conclusions. Data collection, analysis, and interpretation were the responsibility of the authors, independent of artificial intelligence.

## Competing interests

The authors declare no competing interests.

## Notes

### Competing Interest Statement

The authors have declared no competing interest.

### Author Declarations

This study was approved by the Ethics Review Committee of Peking Union Medical College Hospital, Chinese Academy of Medical Sciences, and written informed consent was obtained from all participants (Approval No. JS-3455).

### Summary of Updates

We strengthened the plasma cell analyses by validating the robustness of gene expression programs across whole bone marrow and CD138-enriched samples and by incorporating independent public datasets. We further expanded analysis of post-treatment and residual amyloidogenic plasma cells using paired single-cell V(D)J sequencing and light chain CDR3 sequences for clone tracking. We substantially expanded characterization of the bone marrow immune microenvironment. New analyses include deeper evaluation of interferon responses, prostaglandin signaling, and nonclassical MHC class I signaling. Multiplex immunofluorescence panels were redesigned to localize and quantify prostaglandin pathway proteins and inhibitory receptors in defined plasma cell, myeloid, T cell, and NK cell populations. Flow cytometry was added to validate T cell exhaustion markers and to compare bone marrow/peripheral blood from healthy controls and patients grouped by treatment responses.

