## Supplementary Material for "Single-Cell Analysis Reveals Inflammatory–Immunosuppressive Niches Associated with Suboptimal Response to Daratumumab-Based Therapy in AL Amyloidosis"

#### Table of Contents

Supplementary Fig. S1-S14 Page 2-15

Table S1-S7 Page 16

### Figure S1

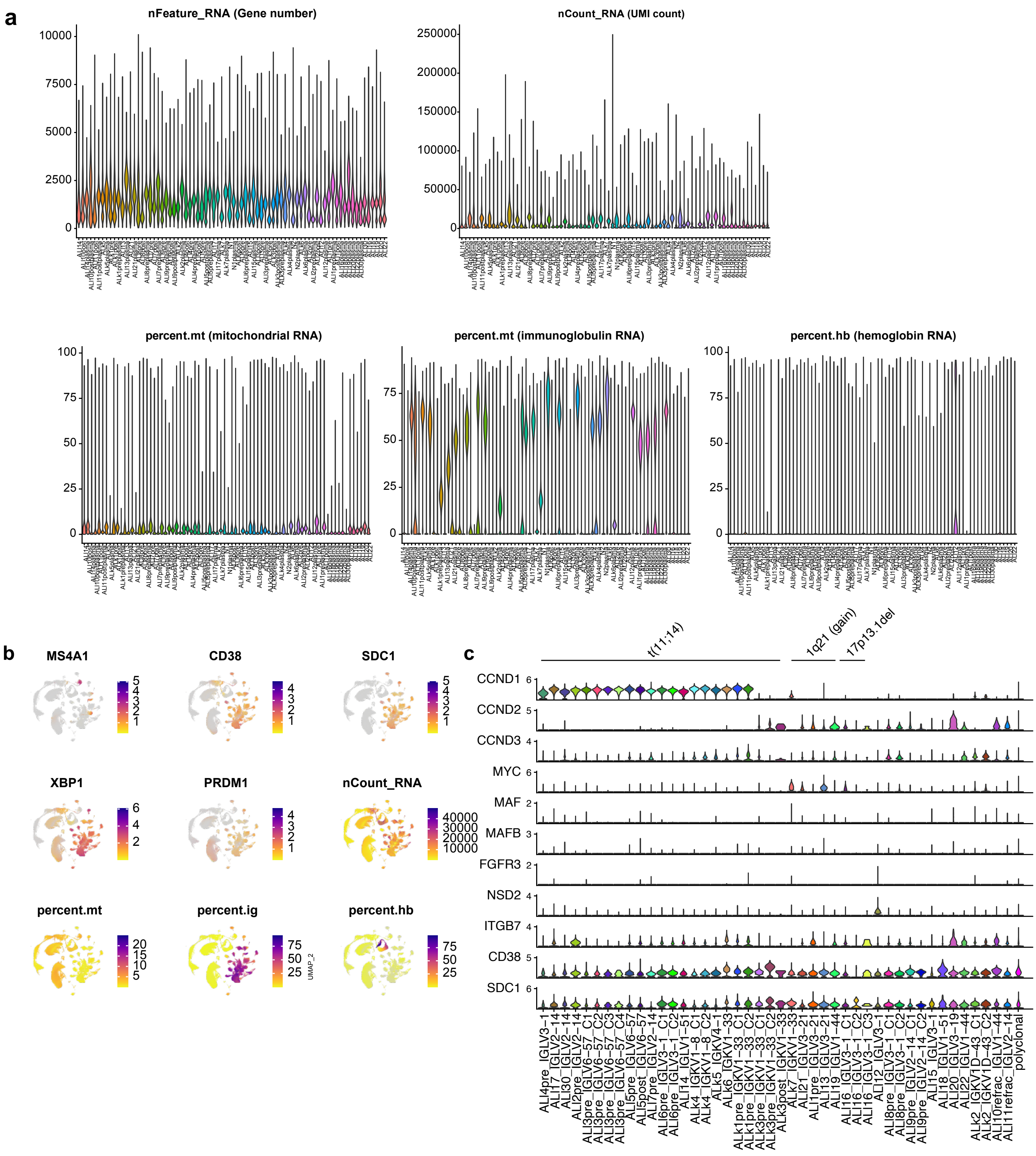
a
nFeature_RNA (Gene number)
nCount_RNA (UMI count)
percent.mt (mitochondrial RNA)
percent.mt (immunoglobulin RNA)
percent.hb (hemoglobin RNA)
b
MS4A1
CD38
SDC1
XBP1
PRDM1
nCount_RNA
percent.mt
percent.ig
percent.hb
UMAP_2
c
t(11;14)
1q21 (gain)
17p13.1del
CCND1
CCND2
CCND3
MYC
MAF
MAFB
FGFR3
NSD2
ITGB7
CD38
SDC1
ALI4pre_IGLV3−1
ALI7_IGLV2−14
ALI30_IGLV2−14
ALI2pre_IGLV2−14
ALI3pre_IGLV6−57_C1
ALI3pre_IGLV6−57_C2
ALI3pre_IGLV6−57_C3
ALI3pre_IGLV6−57_C4
ALI5pre_IGLV6−57
ALI5post_IGLV6−57
ALI7pre_IGLV2−14
ALI6pre_IGLV3−1_C1
ALI6pre_IGLV3−1_C2
ALI14_IGLV1−51
ALk4_IGKV1−8_C1
ALk4_IGKV1−8_C2
ALk5_IGKV4−1
ALk6_IGKV1−33
ALk1pre_IGKV1−33_C1
ALk1pre_IGKV1−33_C2
ALk3pre_IGKV1−33_C1
ALk3pre_IGKV1−33_C2
ALk3post_IGKV1−33
ALk7_IGKV1−33
ALI21_IGLV3−21
ALI1pre_IGLV3−21
ALI13_IGLV3−21
ALI19_IGLV1−44
ALI16_IGLV3−1_C1
ALI16_IGLV3−1_C2
ALI16_IGLV3−1_C3
ALI12_IGLV3−1
ALI8pre_IGLV3−1_C1
ALI8pre_IGLV3−1_C2
ALI9pre_IGLV2−14_C1
ALI9pre_IGLV2−14_C2
ALI15_IGLV3−1
ALI18_IGLV1−51
ALI20_IGLV3−19
ALI22_IGLV1−44
ALk2_IGKV1D−43_C1
ALk2_IGKV1D−43_C2
ALI10refrac_IGLV1−44
ALI11refrac_IGLV2−14
polyclonal

**Supplementary Fig. S1. Quality control metrics.**

**(a)** Violin plots showing gene number per cell (nFeature_RNA), unique molecular identifier (UMI) count per cell (nCount_RNA), fraction of mitochondrial RNA (percent.mt), fraction of immunoglobulin RNA (percent.ig), and fraction of hemoglobin RNA (percent.hb).

**(b)** B cell and plasma cell marker gene expression and quality-control metrics.

**(c)** Violin plots showing the expression of genes commonly upregulated in plasma cell diseases, aligned with cytogenetic abnormalities

### Figure S2

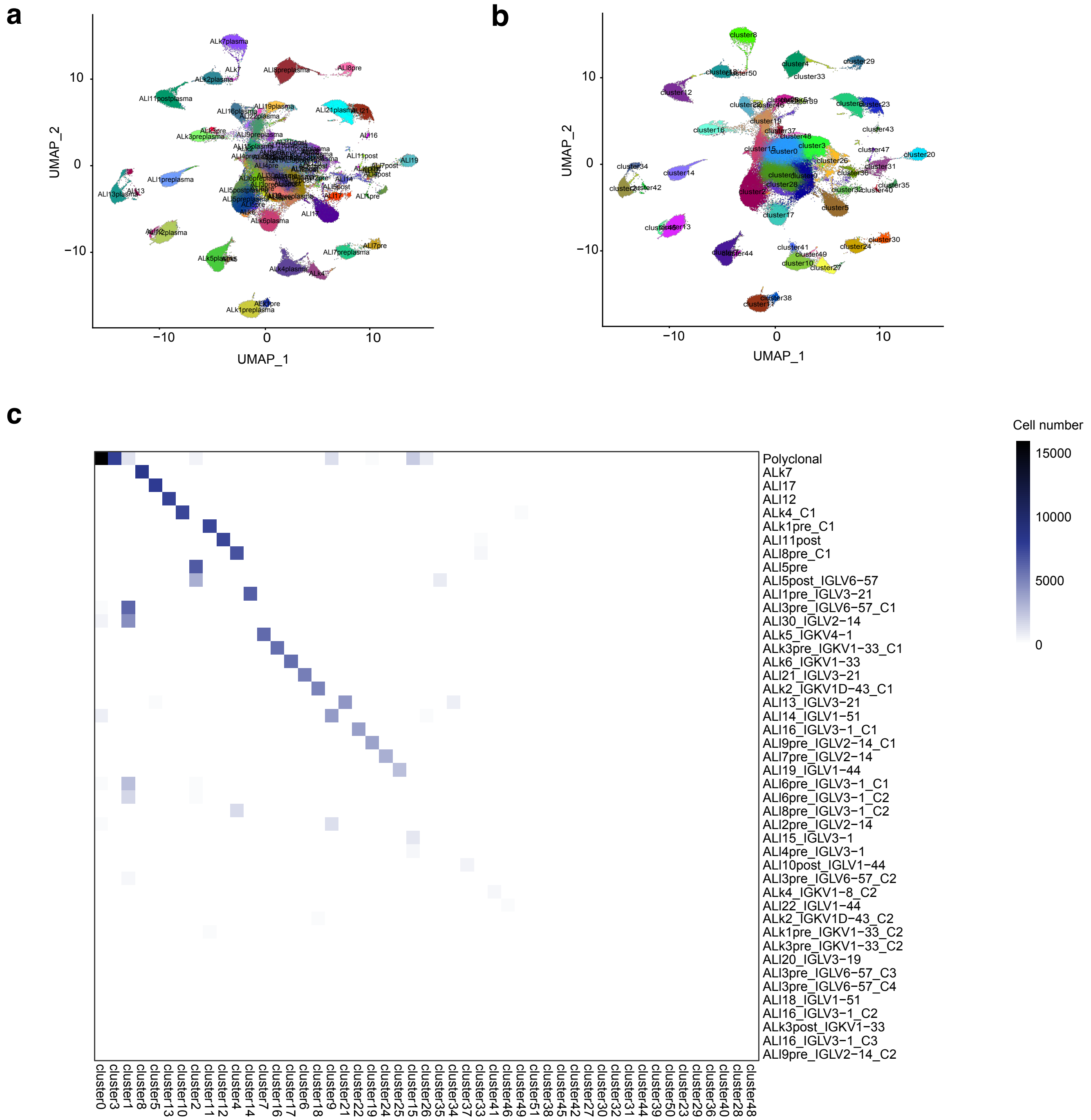

a
b
c
UMAP_1
UMAP_2
10
0
−10
Cell number
15000
10000
5000
0
Polyclonal
ALk7
ALl17
ALl12
ALk4_C1
ALk1pre_C1
ALl11post
ALl8pre_C1
ALl5pre
ALl5post_IGLV6−57
ALl1pre_IGLV3−21
ALl3pre_IGLV6−57_C1
ALl30_IGLV2−14
ALk5_IGKV4−1
ALk3pre_IGKV1−33_C1
ALk6_IGKV1−33
ALl21_IGLV3−21
ALk2_IGKV1D−43_C1
ALl13_IGLV3−21
ALl14_IGLV1−51
ALl16_IGLV3−1_C1
ALl9pre_IGLV2−14_C1
ALl7pre_IGLV2−14
ALl19_IGLV1−44
ALl6pre_IGLV3−1_C1
ALl6pre_IGLV3−1_C2
ALl8pre_IGLV3−1_C2
ALl2pre_IGLV2−14
ALl15_IGLV3−1
ALl4pre_IGLV3−1
ALl10post_IGLV1−44
ALl3pre_IGLV6−57_C2
ALk4_IGKV1−8_C2
ALl22_IGLV1−44
ALk2_IGKV1D−43_C2
ALk1pre_IGKV1−33_C2
ALk3pre_IGKV1−33_C2
ALl20_IGLV3−19
ALl3pre_IGLV6−57_C3
ALl3pre_IGLV6−57_C4
ALl18_IGLV1−51
ALl16_IGLV3−1_C2
ALk3post_IGKV1−33
ALl16_IGLV3−1_C3
ALl9pre_IGLV2−14_C2
cluster0
cluster3
cluster1
cluster8
cluster5
cluster13
cluster10
cluster2
cluster11
cluster12
cluster4
cluster14
cluster7
cluster16
cluster17
cluster6
cluster18
cluster9
cluster21
cluster22
cluster19
cluster24
cluster25
cluster15
cluster26
cluster35
cluster34
cluster37
cluster33
cluster41
cluster46
cluster49
cluster51
cluster38
cluster45
cluster42
cluster27
cluster30
cluster32
cluster31
cluster44
cluster39
cluster50
cluster23
cluster29
cluster36
cluster40
cluster28
cluster48

**Supplementary Fig. S2. Plasma cell clonality.**

**(a)** Uniform manifold approximation and projection (UMAP) visualization of all bone marrow plasma cells grouped by sample ID.

**(b)** UMAP visualization of all bone marrow cells with k-nearest neighbour graph–based Louvain clustering.

**(c)** Heatmap showing the number of plasma cells per clonotype assigned to each cluster.

### Figure S3

**a**

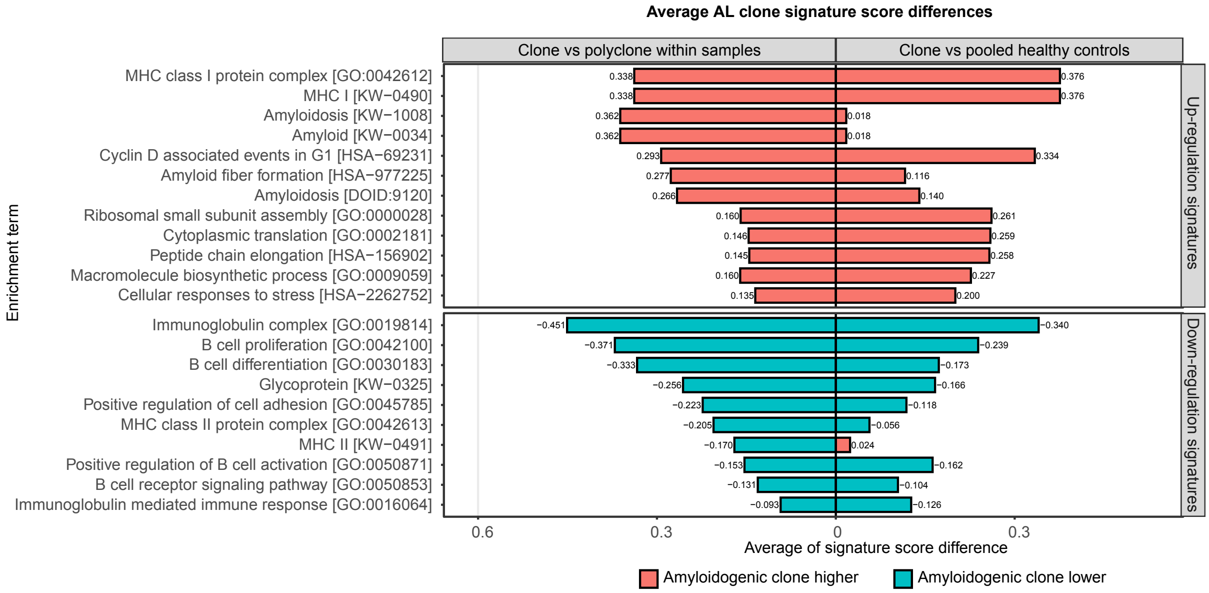


(Gort-Freitas et al., Blood, 2025)

**b**

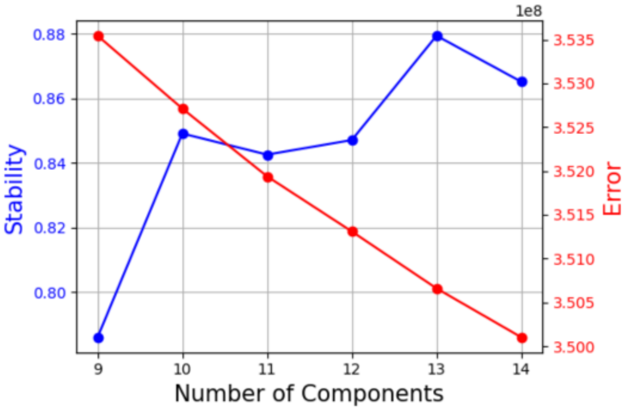

**Supplementary Fig. S3. Gene expression program inference.**

**(a)** Bar plot showing differences in amyloidogenic plasma cell signature scores between amyloidogenic plasma cell clones and polyclonal plasma cells (Dataset: Gort-Freitas et al., Blood, 2025).

**(b)** Stability–component number–error rate plot identifying the optimized number of components in consensus non-negative matrix factorization (cNMF) analysis.

#### Figure S4

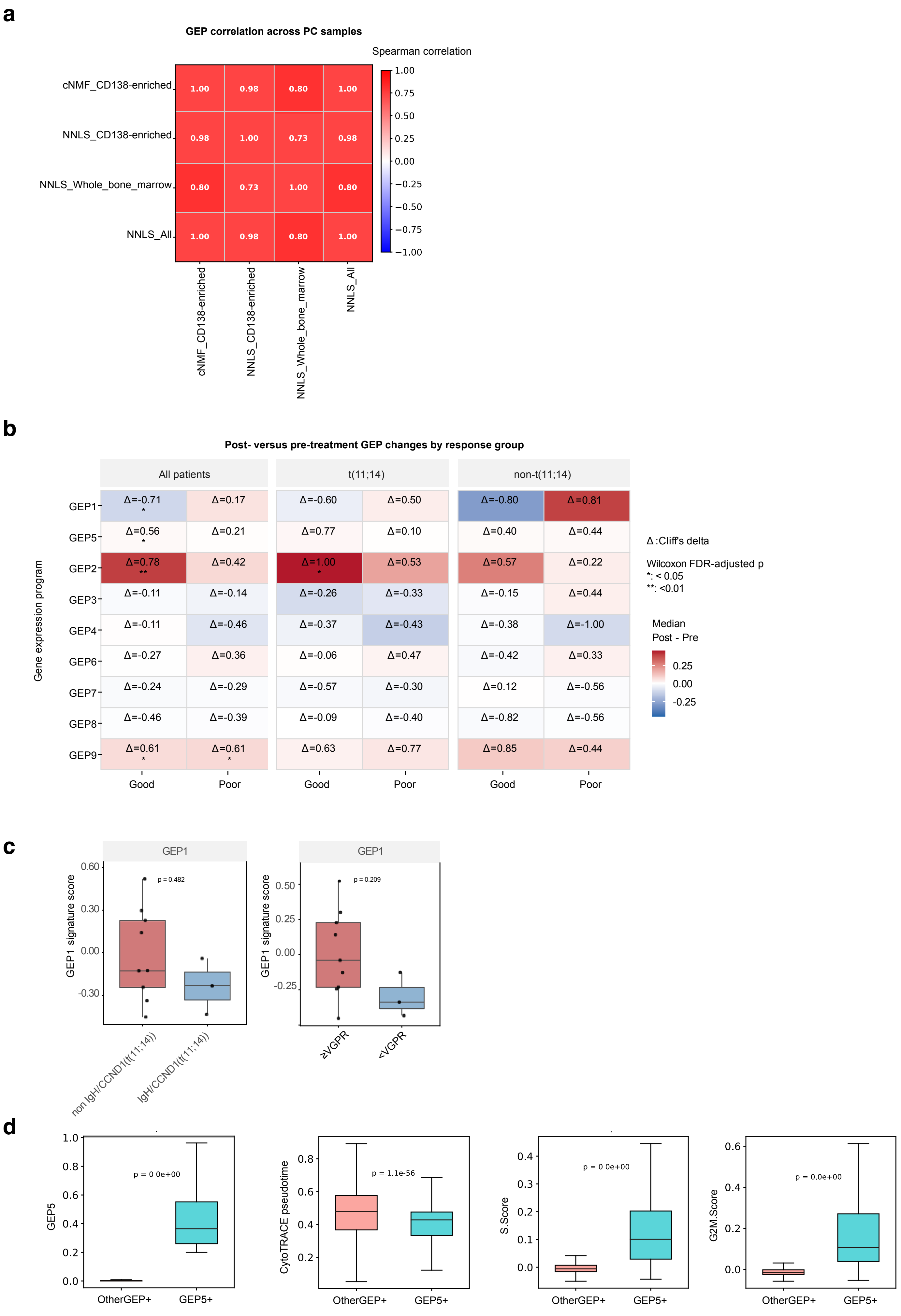
a
GEP correlation across PC samples
Spearman correlation
cNMF_CD138-enriched
NNLS_CD138-enriched
NNLS_Whole_bone_marrow
NNLS_All
b
Post- versus pre-treatment GEP changes by response group
All patients
t(11;14)
non-t(11;14)
Gene expression program
Good
Poor
Δ:Cliff's delta
Wilcoxon FDR-adjusted p
*: < 0.05
**: <0.01
Median
Post - Pre
c
GEP1
GEP1 signature score
p = 0.482
p = 0.209
non IgH/CCND1(t(11;14))
IgH/CCND1(t(11;14))
≥VGPR
<VGPR
d
GEP5
CytoTRACE pseudotime
S.Score
G2M.Score
p = 0.0e+00
p = 1.1e-56
OtherGEP+
GEP5+

**Supplementary Fig. S4 Gene expression program scores.**

**(a)** Spearman correlation of plasma-cell gene expression program (GEP) scores between whole bone marrow and CD138-enriched samples.

**(b)** Heatmap showing the post-vs-pre GEP score changes in all patients, patients with t(11;14), and patients without t(11;14).

**(c)** Bulk RNA-seq validation of GEP1 score differences between good vs suboptimal response groups and between non-t(11;14) vs t(11;14) cytogenetic subgroups.

**(d)** Inferred stemness (CytoTRACE pseudotime) and cell cycle scores (G2M.Score and S.Score) in the mitotic plasma cell state (GEP5+) versus other cell states.

### Figure S5

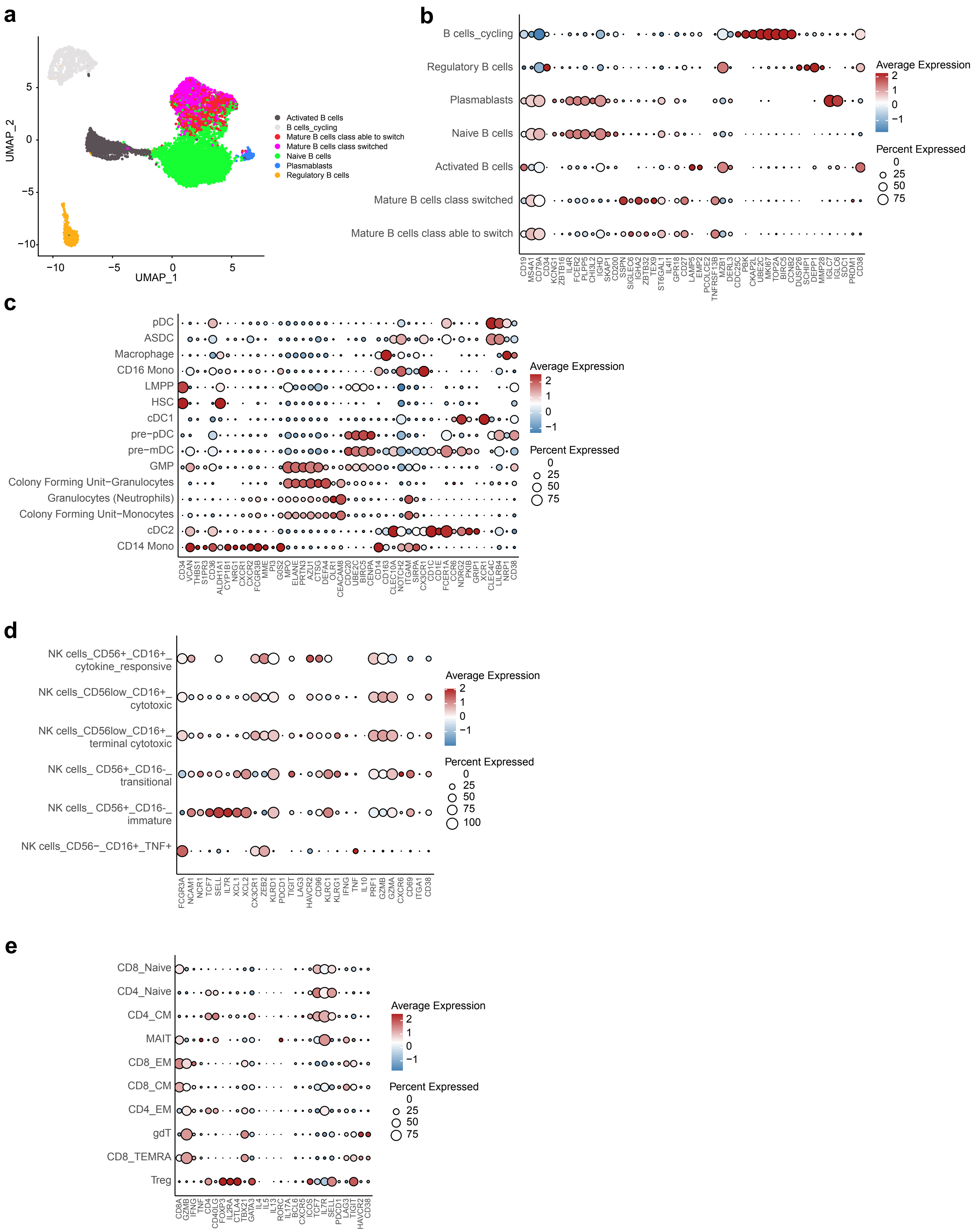
a
b
c
d
e
UMAP_1
UMAP_2
Activated B cells
B cells_cycling
Mature B cells class able to switch
Mature B cells class switched
Naive B cells
Plasmablasts
Regulatory B cells
Average Expression
Percent Expressed

**Supplementary Fig. S5. Immune cell marker gene expression.**

**(a)** Uniform manifold approximation and projection (UMAP) visualization of all B cell subsets, annotated manually.

**(b)** Dot plot showing expression of B cell subtype marker genes, annotated manually.

**(c)** Dot plot showing expression of myeloid lineage marker genes, annotated by Azimuth.

**(d)** Dot plot showing expression of NK cell subtype marker genes, annotated manually.

**(e)** Dot plot showing expression of T cell subtype marker genes, annotated by starCAT.

### Figure S6

**a**

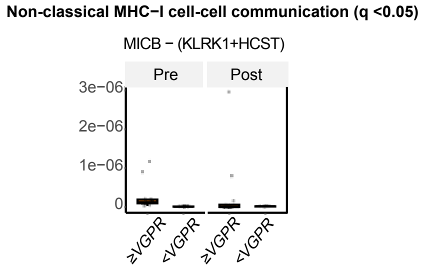
Non-classical MHC−I cell-cell communication (q <0.05)
MICB − (KLRK1+HCST)
Pre
Post
3e−06
2e−06
1e−06
0
≥VGPR
<VGPR
≥VGPR
<VGPR

**Supplementary Fig. S6. CellChat cell-cell communication.**

**(a)** Non-classical MHC class I signaling ligand-receptor pairs, *MICB*–(*KLRK1+HCST*), associated with hematologic response.

### Figure S7

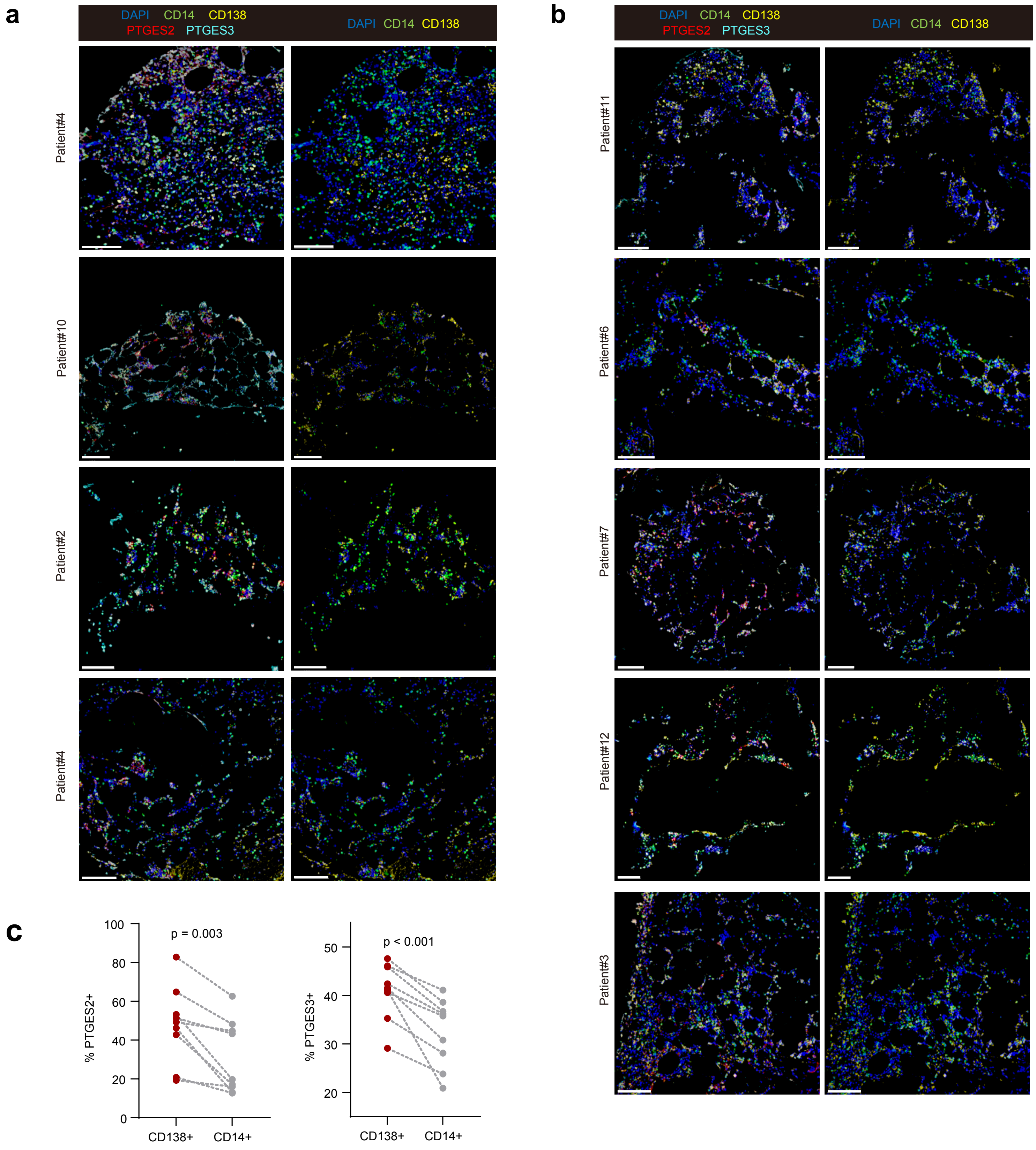
a
DAPI CD14 CD138
PTGES2 PTGES3
DAPI CD14 CD138
Patient#4
Patient#10
Patient#2
Patient#4
b
DAPI CD14 CD138
PTGES2 PTGES3
DAPI CD14 CD138
Patient#11
Patient#6
Patient#7
Patient#12
Patient#3
c
p = 0.003
% PTGES2+
100
80
60
40
20
0
CD138+
CD14+
p < 0.001
% PTGES3+
50
40
30
20
CD138+
CD14+

**Supplementary Fig. S7. Multiplex immunofluorescence.**

**(a)** Representative multiplex immunofluorescence images of bone marrow sections from suboptimal responders, stained for DAPI, CD14, CD138, PTGES2, and PTGES3. Scale bars: 100 μm.

**(b)** Representative multiplex immunofluorescence images of bone marrow sections from good responders, stained for DAPI, CD14, CD138, PTGES2, and PTGES3. Scale bars: 100 μm.

**(c)** Percentages of PTGES2$^{+}$ or PTGES3$^{+}$ cells among CD138$^{+}$ and CD14$^{+}$ cell populations in all bone marrow samples (n = 8 per group).

### Figure S8

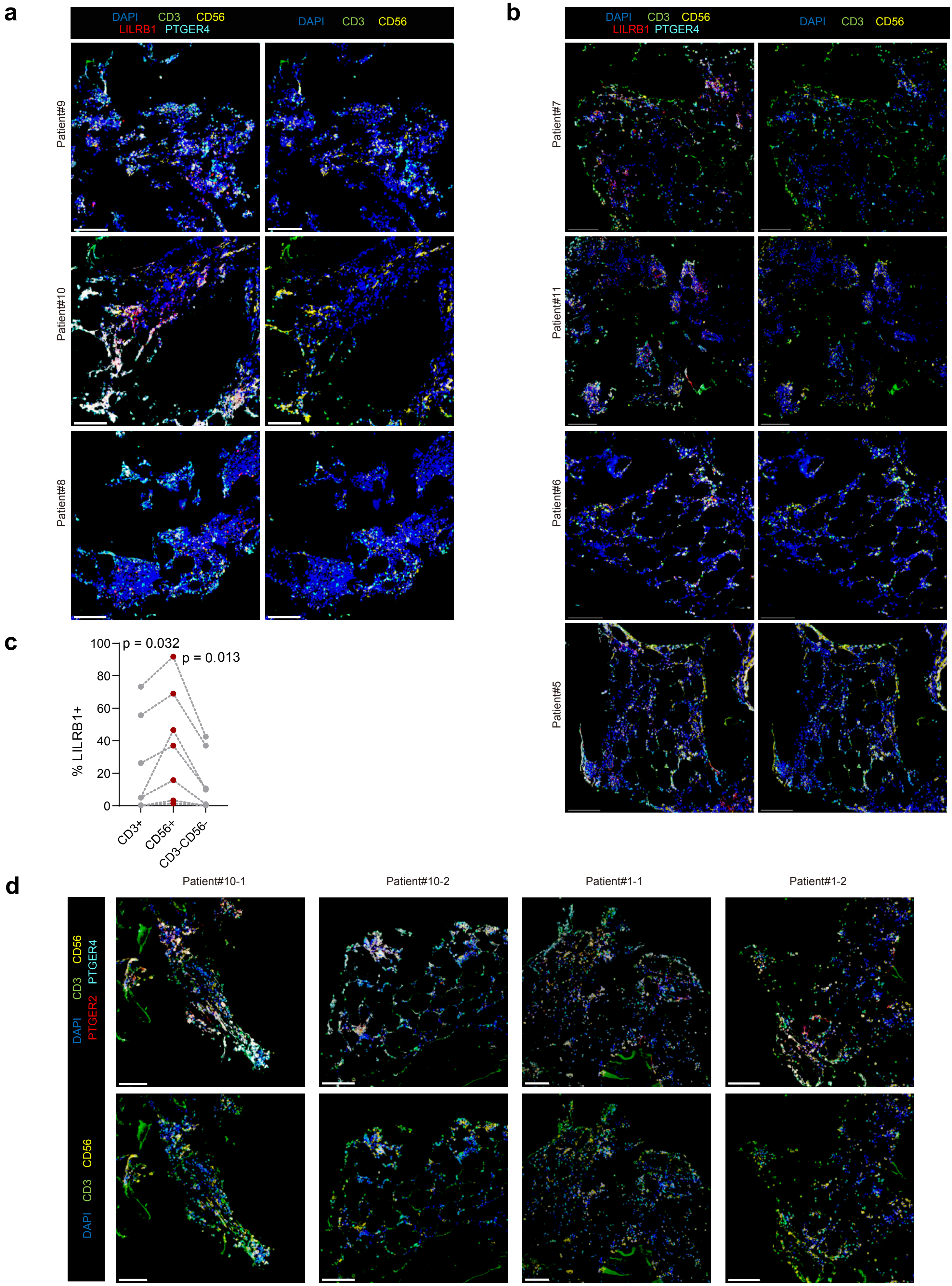

a
DAPI CD3 CD56
LILRB1 PTGER4
DAPI CD3 CD56
Patient#9
Patient#10
Patient#8
b
DAPI CD3 CD56
LILRB1 PTGER4
DAPI CD3 CD56
Patient#7
Patient#11
Patient#6
Patient#5
c
p = 0.032
p = 0.013
% LILRB1+
100
80
60
40
20
0
CD3+
CD56+
CD3-CD56-
d
Patient#10-1
Patient#10-2
Patient#1-1
Patient#1-2
DAPI CD3 CD56
PTGER2 PTGER4
DAPI CD3 CD56

**Supplementary Fig. S8. Multiplex immunofluorescence.**

**(a)** Representative multiplex immunofluorescence images of bone marrow sections from suboptimal responders, stained for DAPI, CD3, CD56, LILRB1, and PTGER4. Scale bars: 100 μm.

**(b)** Representative multiplex immunofluorescence images of bone marrow sections from good responders, stained for DAPI, CD3, CD56, LILRB1, and PTGER4. Scale bars: 100 μm.

**(c)** Percentages of $LILRB1^{+}PTGER4^{+}$ cells among $CD3^{+}$, $CD56^{+}$, and $CD3^{-}CD56^{-}$ cell populations in all bone marrow samples (n = 7 per group).

**(d)** Representative multiplex immunofluorescence images of bone marrow sections from suboptimal responders, stained for DAPI, CD3, CD56, PTGER2, and PTGER4. Scale bars: 100 μm.

**Figure S9**

**a**

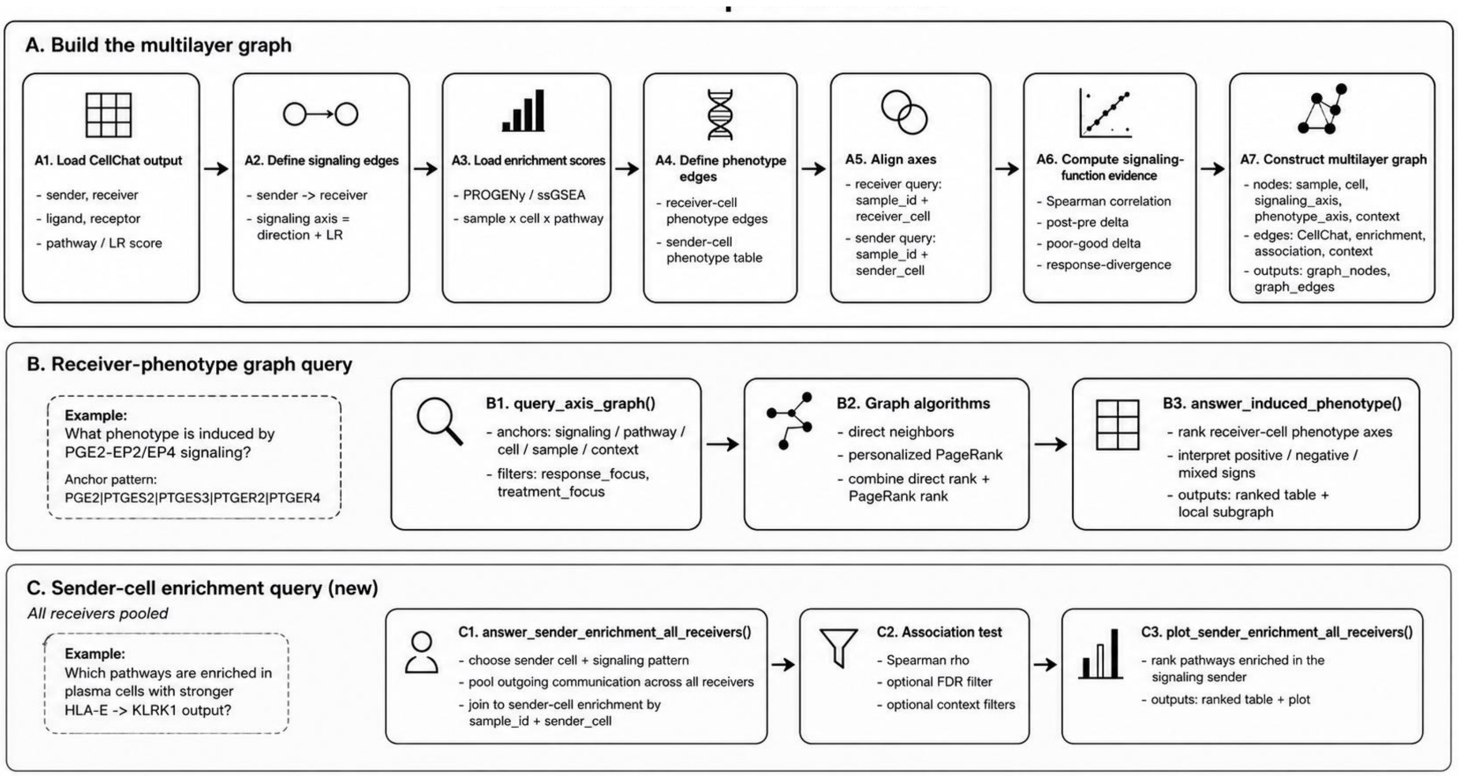
A. Build the multilayer graph
A1. Load CellChat output
- sender, receiver
- ligand, receptor
- pathway / LR score
A2. Define signaling edges
- sender -> receiver
- signaling axis = direction + LR
A3. Load enrichment scores
- PROGENy / ssGSEA
- sample x cell x pathway
A4. Define phenotype edges
- receiver-cell phenotype edges
- sender-cell phenotype table
A5. Align axes
- receiver query: sample_id + receiver_cell
- sender query: sample_id + sender_cell
A6. Compute signaling-function evidence
- Spearman correlation
- post-pre delta
- poor-good delta
- response-divergence
A7. Construct multilayer graph
- nodes: sample, cell, signaling_axis, phenotype_axis, context
- edges: CellChat, enrichment, association, context
- outputs: graph_nodes, graph_edges
B. Receiver-phenotype graph query
Example:
What phenotype is induced by PGE2-EP2/EP4 signaling?
Anchor pattern:
PGE2|PTGES2|PTGES3|PTGER2|PTGER4
B1. query_axis_graph()
- anchors: signaling / pathway / cell / sample / context
- filters: response_focus, treatment_focus
B2. Graph algorithms
- direct neighbors
- personalized PageRank
- combine direct rank + PageRank rank
B3. answer_induced_phenotype()
- rank receiver-cell phenotype axes
- interpret positive / negative / mixed signs
- outputs: ranked table + local subgraph
C. Sender-cell enrichment query (new)
All receivers pooled
Example:
Which pathways are enriched in plasma cells with stronger HLA-E -> KLRK1 output?
C1. answer_sender_enrichment_all_receivers()
- choose sender cell + signaling pattern
- pool outgoing communication across all receivers
- join to sender-cell enrichment by sample_id + sender_cell
C2. Association test
- Spearman rho
- optional FDR filter
- optional context filters
C3. plot_sender_enrichment_all_receivers()
- rank pathways enriched in the signaling sender
- outputs: ranked table + plot

**Supplementary Fig. S9. ALNicheGraph.**

**(a)** Overview of ALNicheGraph workflow.

### Figure S10

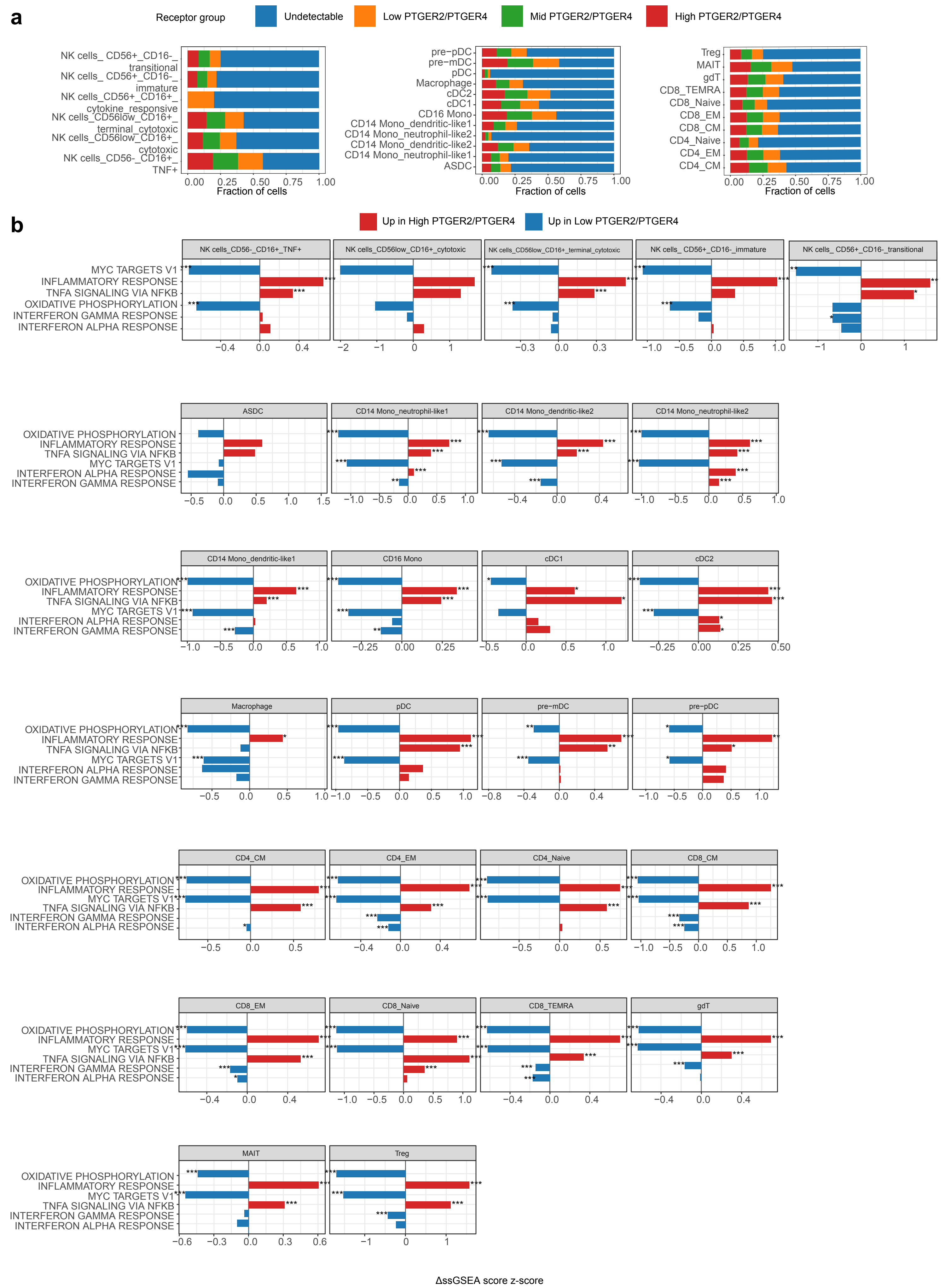

a
Receptor group
Undetectable
Low PTGER2/PTGER4
Mid PTGER2/PTGER4
High PTGER2/PTGER4
NK cells_ CD56+_CD16-_transitional
NK cells_ CD56+_CD16-_immature
NK cells_CD56+_CD16+_cytokine_responsive
NK cells_CD56low_CD16+_terminal_cytotoxic
NK cells_CD56low_CD16+_cytotoxic
NK cells_CD56-_CD16+_TNF+
pre−pDC
pre−mDC
pDC
Macrophage
cDC2
cDC1
CD16 Mono
CD14 Mono_dendritic-like1
CD14 Mono_neutrophil-like2
CD14 Mono_dendritic-like2
CD14 Mono_neutrophil-like1
ASDC
Treg
MAIT
gdT
CD8_TEMRA
CD8_Naive
CD8_EM
CD8_CM
CD4_Naive
CD4_EM
CD4_CM
Fraction of cells
b
Up in High PTGER2/PTGER4
Up in Low PTGER2/PTGER4
NK cells_CD56-_CD16+_TNF+
NK cells_CD56low_CD16+_cytotoxic
NK cells_CD56low_CD16+_terminal_cytotoxic
NK cells_ CD56+_CD16-_immature
NK cells_ CD56+_CD16-_transitional
ASDC
CD14 Mono_neutrophil-like1
CD14 Mono_dendritic-like2
CD14 Mono_neutrophil-like2
CD14 Mono_dendritic-like1
CD16 Mono
cDC1
cDC2
Macrophage
pDC
pre−mDC
pre−pDC
CD4_CM
CD4_EM
CD4_Naive
CD8_CM
CD8_EM
CD8_Naive
CD8_TEMRA
gdT
MAIT
Treg
MYC TARGETS V1
INFLAMMATORY RESPONSE
TNFA SIGNALING VIA NFKB
OXIDATIVE PHOSPHORYLATION
INTERFERON GAMMA RESPONSE
INTERFERON ALPHA RESPONSE
ΔssGSEA score z-score

**Supplementary Fig. S10. Interferon response and *PTGER2/PTGER4* expression.**

**(a)** NK/myeloid/T cells grouped by *PTGER4/PTGER2* expression level.

**(b)** Differential enrichment of functional pathways in *PTGER4/PTGER2*$^{high}$ versus *PTGER4/PTGER2*$^{low}$ immune cells.

### Figure S11

**a**

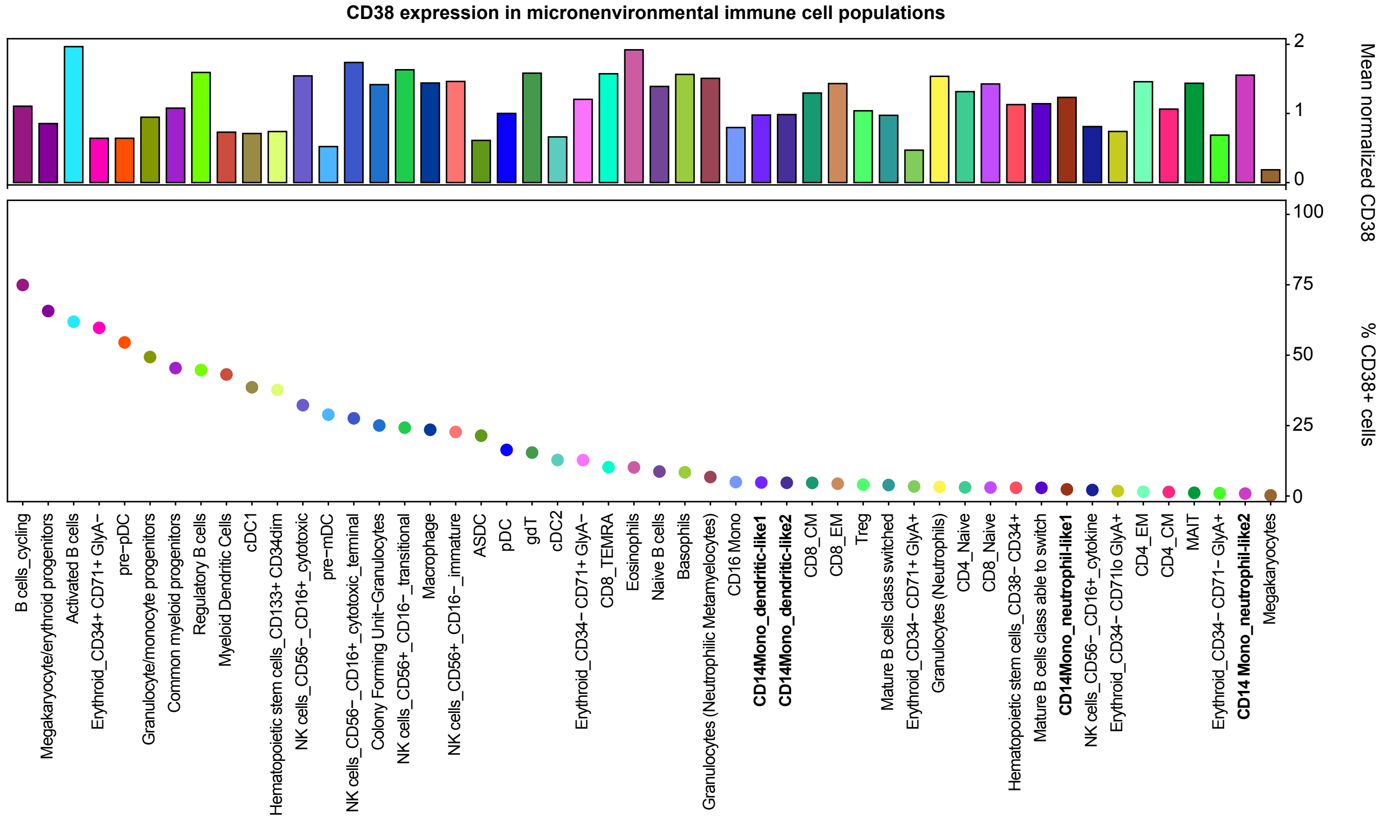

CD38 expression in micronenvironmental immune cell populations
Mean normalized CD38
% CD38+ cells
0
1
2
0
25
50
75
100
B cells_cycling
Megakaryocyte/erythroid progenitors
Activated B cells
Erythroid_CD34+ CD71+ GlyA−
pre−pDC
Granulocyte/monocyte progenitors
Common myeloid progenitors
Regulatory B cells
Myeloid Dendritic Cells
cDC1
Hematopoietic stem cells_CD133+ CD34dim
NK cells_CD56−_CD16+_cytotoxic
pre−mDC
NK cells_CD56−_CD16+ cytotoxic_terminal
Colony Forming Unit−Granulocytes
NK cells_CD56+_CD16−_transitional
Macrophage
NK cells_CD56+_CD16−_immature
ASDC
pDC
gdT
cDC2
Erythroid_CD34− CD71+ GlyA−
CD8_TEMRA
Eosinophils
Naive B cells
Basophils
Granulocytes (Neutrophilic Metamyelocytes)
CD16 Mono
CD14Mono_dendritic-like1
CD14Mono_dendritic-like2
CD8_CM
CD8_EM
Treg
Mature B cells class switched
Erythroid_CD34− CD71+ GlyA+
Granulocytes (Neutrophils)
CD4_Naive
CD8_Naive
Hematopoietic stem cells_CD38− CD34+
Mature B cells class able to switch
CD14Mono_neutrophil-like1
NK cells_CD56−_CD16+_cytokine
Erythroid_CD34− CD71lo GlyA+
CD4_EM
CD4_CM
MAIT
Erythroid_CD34− CD71− GlyA+
CD14 Mono_neutrophil-like2
Megakaryocytes

**Supplementary Fig. S11. *CD38* expression in immune cell populations.**

**(a)** *CD38* expression across bone marrow immune cell populations. The upper bar plot shows the mean normalized *CD38* expression within each cell population, whereas the lower dot plot shows the percentage of $CD38^+$ cells in the corresponding cell population.

### Figure S12

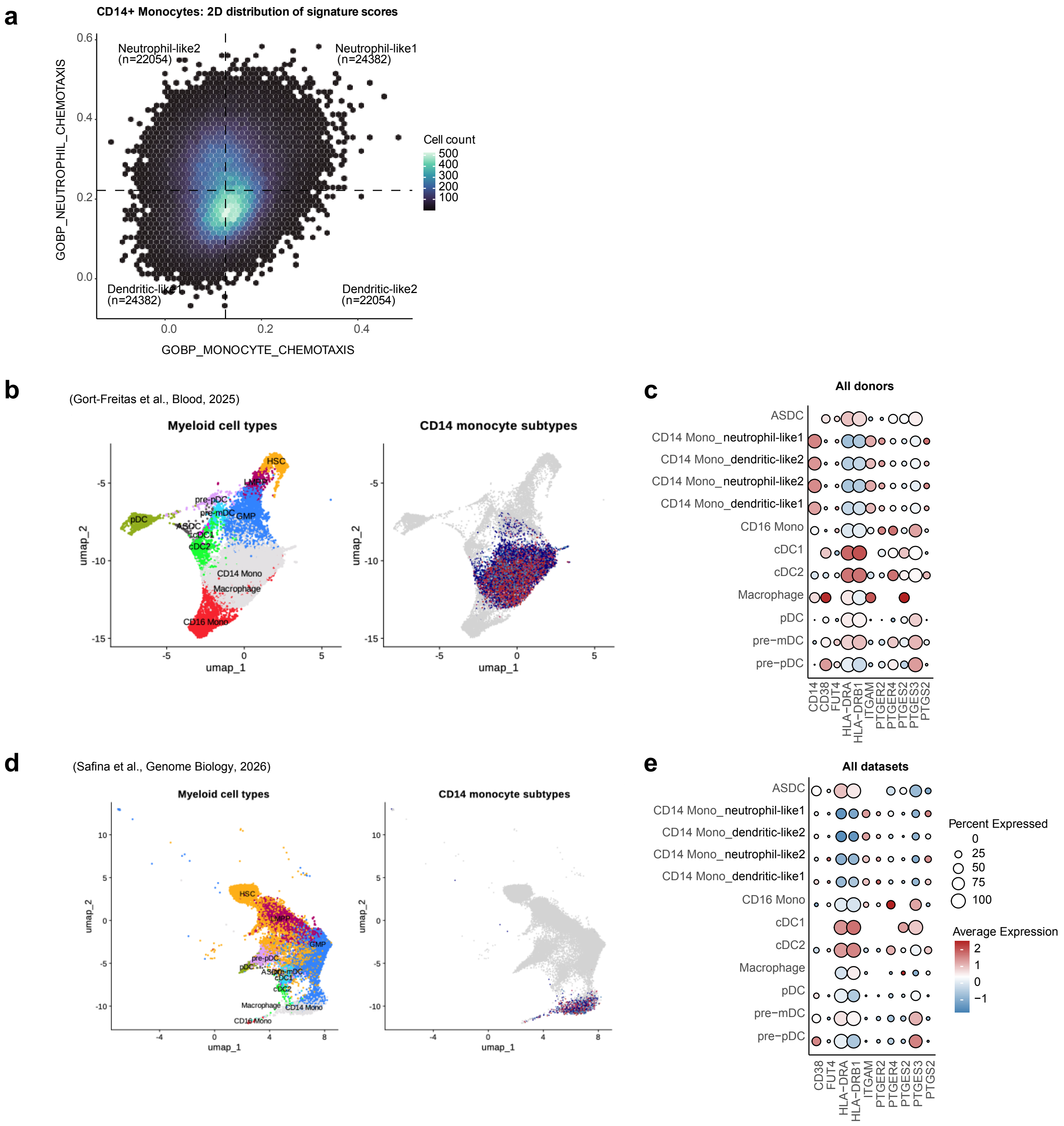
a
CD14+ Monocytes: 2D distribution of signature scores
Neutrophil-like2 (n=22054)
Neutrophil-like1 (n=24382)
Dendritic-like1 (n=24382)
Dendritic-like2 (n=22054)
GOBP_NEUTROPHIL_CHEMOTAXIS
GOBP_MONOCYTE_CHEMOTAXIS
Cell count
500
400
300
200
100
b
(Gort-Freitas et al., Blood, 2025)
Myeloid cell types
CD14 monocyte subtypes
HSC
LMPP
pre-pDC
pre-mDC
GMP
pDC
ASDC
cDC1
cDC2
CD14 Mono
Macrophage
CD16 Mono
umap_1
umap_2
c
All donors
ASDC
CD14 Mono_neutrophil-like1
CD14 Mono_dendritic-like2
CD14 Mono_neutrophil-like2
CD14 Mono_dendritic-like1
CD16 Mono
cDC1
cDC2
Macrophage
pDC
pre−mDC
pre−pDC
CD14
CD38
FUT4
HLA−DRA
HLA−DRB1
ITGAM
PTGER2
PTGER4
PTGES2
PTGES3
PTGS2
d
(Safina et al., Genome Biology, 2026)
Myeloid cell types
CD14 monocyte subtypes
umap_1
umap_2
e
All datasets
CD38
FUT4
HLA−DRA
HLA−DRB1
ITGAM
PTGER2
PTGER4
PTGES2
PTGES3
PTGS2
Percent Expressed
0
25
50
75
100
Average Expression
2
1
0
−1

**Supplementary Fig. S12. Monocyte subsets and gene expression.**

**(a)** Expression of monocyte chemotaxis and neutrophil chemotaxis signatures in single cells.

**(b)** Myeloid cell subsets in patients with systemic light-chain amyloidosis (AL) and healthy donors (Dataset: Gort-Freitas et al., Blood, 2025).

**(c)** Myeloid cell marker gene expression in patients with AL and healthy donors (Dataset: Gort-Freitas et al., Blood, 2025).

**(d)** Myeloid cell subsets from 98 healthy donors (Dataset: Safina et al., Genome Biology, 2026).

**(e)** Myeloid cell marker gene expression in 98 healthy donors (Dataset: Safina et al., Genome Biology, 2026).

### Figure S13

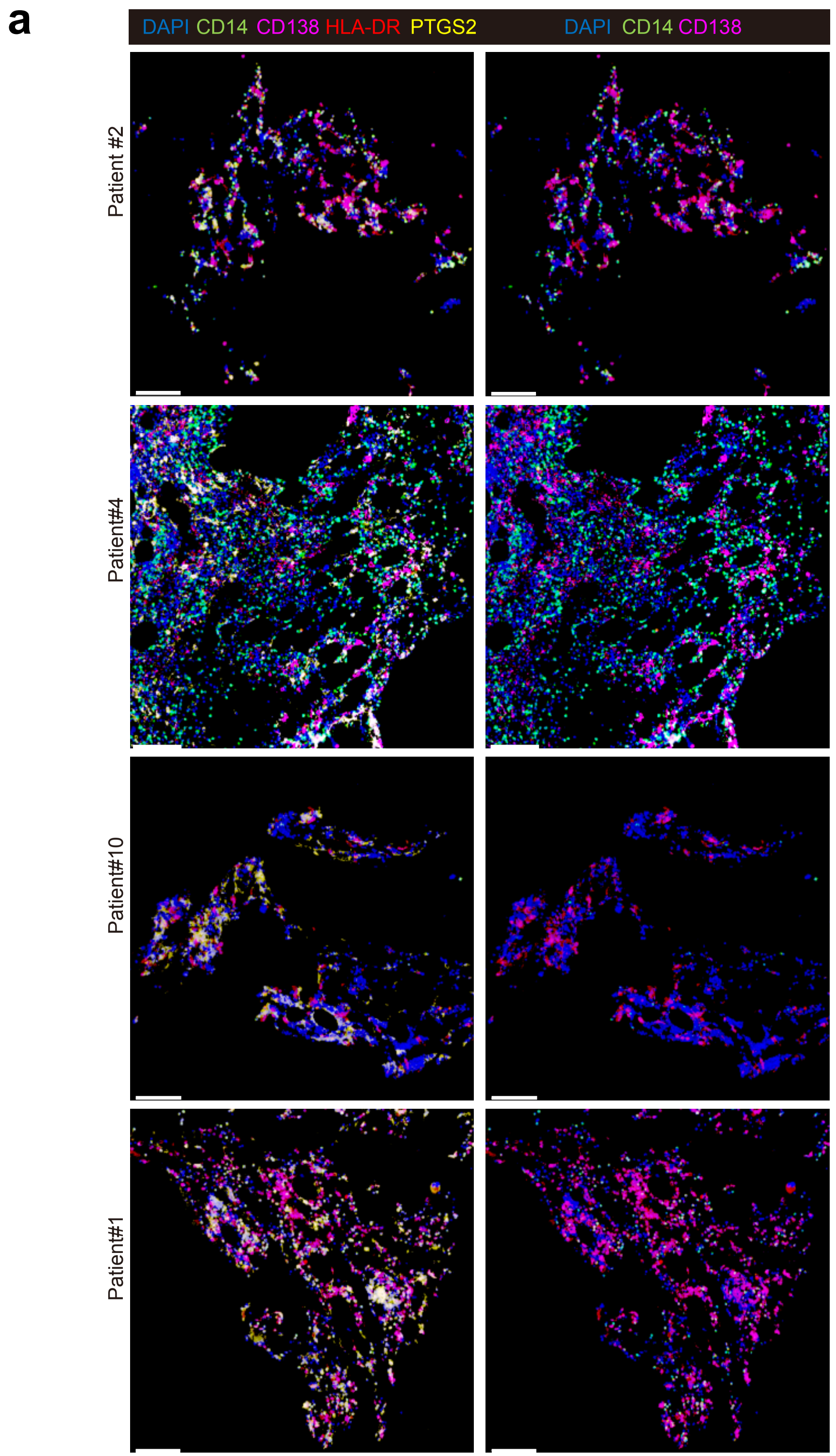
a
DAPI CD14 CD138 HLA-DR PTGS2
DAPI CD14 CD138
Patient #2
Patient#4
Patient#10
Patient#1

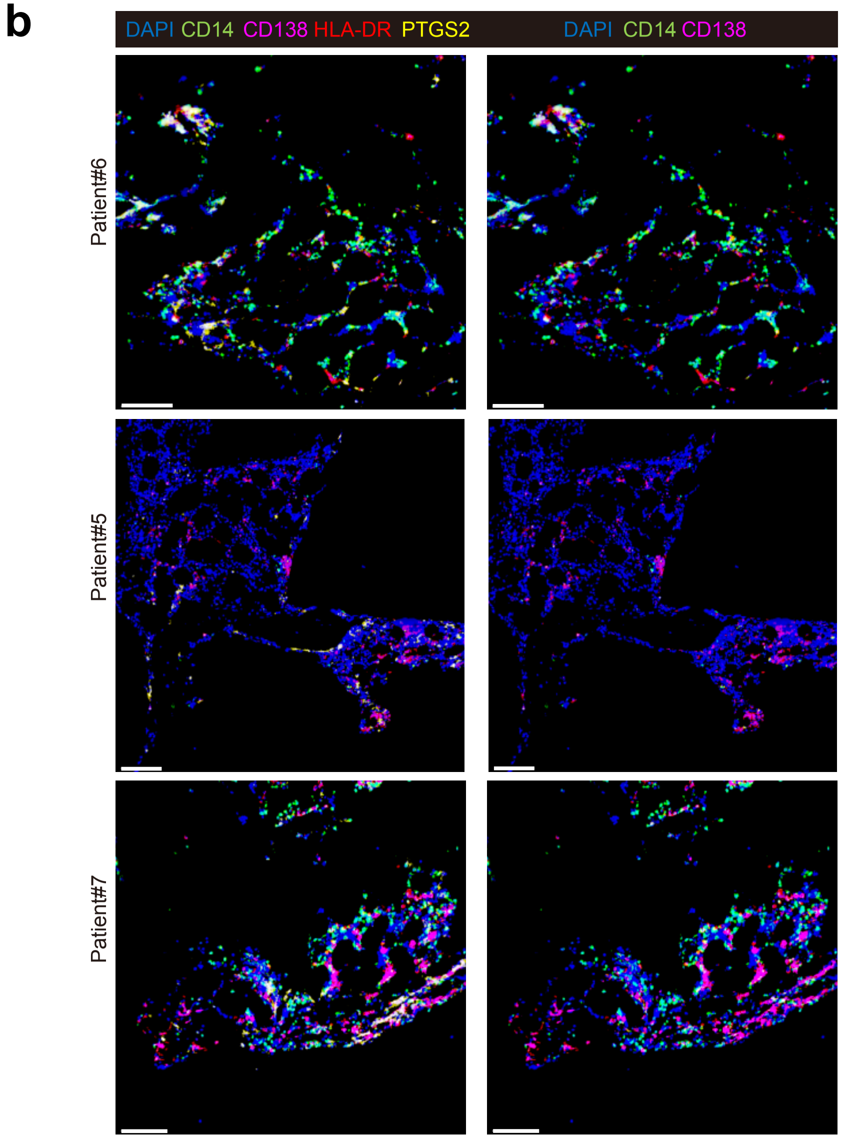
b
DAPI CD14 CD138 HLA-DR PTGS2
DAPI CD14 CD138
Patient#6
Patient#5
Patient#7

**Supplementary Fig. S13. Multiplex immunofluorescence.**

**(a)** Representative multiplex immunofluorescence images of bone marrow sections from suboptimal responders, stained for DAPI, CD14, CD138, HLA-DR and PTGS2. Scale bars: 100 µm.

**(b)** Representative multiplex immunofluorescence images of bone marrow sections from good responders, stained for DAPI, CD14, CD138, HLA-DR and PTGS2. Scale bars: 100 µm.

### Figure S14

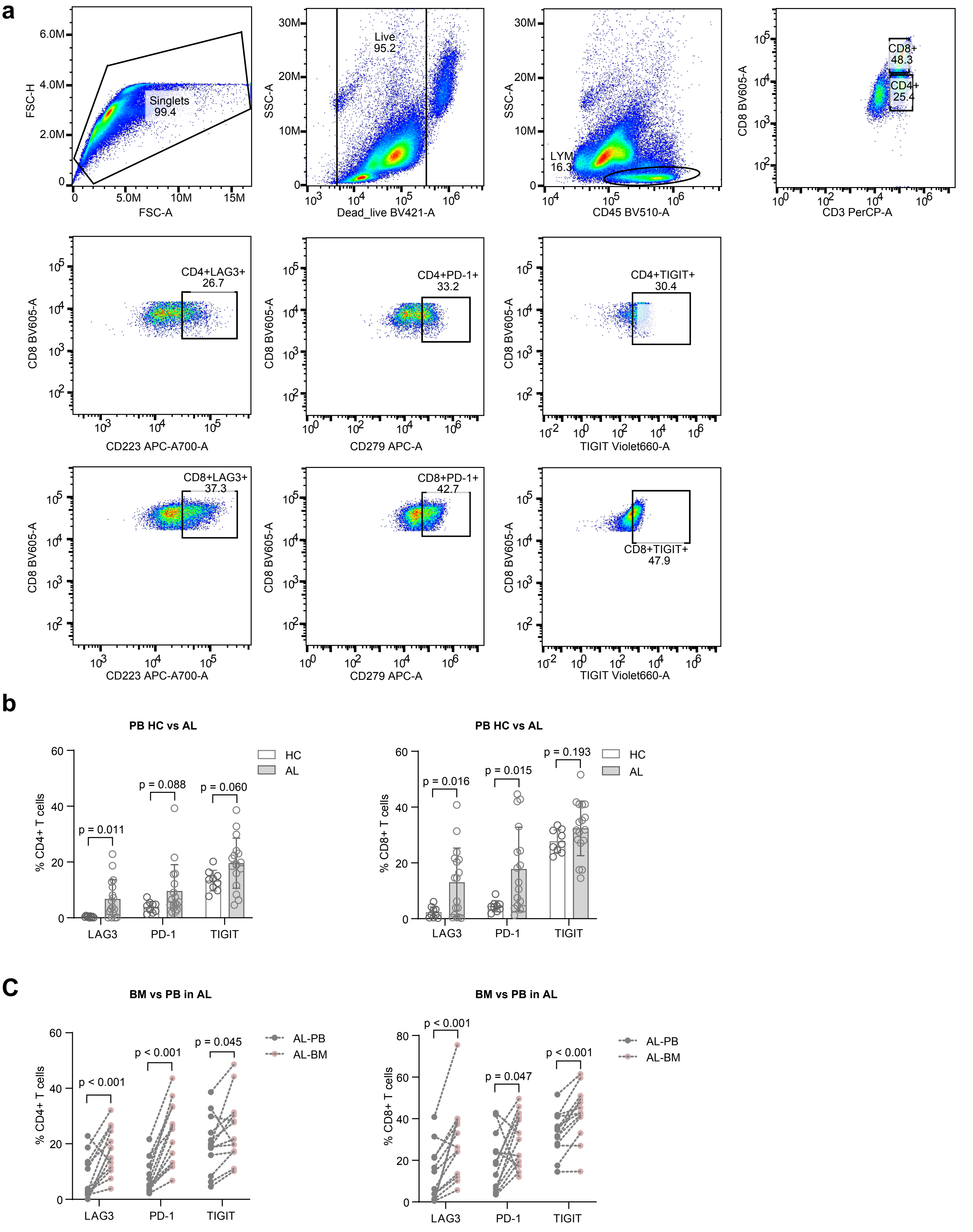

a
Singlets
99.4
FSC-H
FSC-A
Live
95.2
SSC-A
Dead_live BV421-A
LYM
16.3
CD45 BV510-A
CD8+
48.3
CD4+
25.4
CD8 BV605-A
CD3 PerCP-A
CD4+LAG3+
26.7
CD223 APC-A700-A
CD4+PD-1+
33.2
CD279 APC-A
CD4+TIGIT+
30.4
TIGIT Violet660-A
CD8+LAG3+
37.3
CD8+PD-1+
42.7
CD8+TIGIT+
47.9
b
PB HC vs AL
HC
AL
% CD4+ T cells
p = 0.011
p = 0.088
p = 0.060
LAG3
PD-1
TIGIT
% CD8+ T cells
p = 0.016
p = 0.015
p = 0.193
C
BM vs PB in AL
AL-PB
AL-BM
p < 0.001
p < 0.001
p = 0.045
p < 0.001
p = 0.047
p < 0.001

**Supplementary Fig. S14. Flow cytometry for T-cell exhaustion.**

**(a)** Representative flow cytometric gating strategy of bone marrow mononuclear cells (Patient #5).

**(b)** Percentages of peripheral blood $CD4^+$ and $CD8^+$ T cells positive for the exhaustion markers, measured by flow cytometry, in healthy controls (n = 9) and patients with systemic light-chain amyloidosis (AL) (n = 17).

**(c)** Baseline percentages of $CD4^+$ and $CD8^+$ T cells positive for the exhaustion markers in bone marrow and peripheral blood in patients with AL (n = 13 per group), measured by flow cytometry.

**Table S1. De-identified clinical characteristics of patients in the single-cell cohort.**

Please see the attached file in Source Data.

**Table S2. Top 100 genes for each gene expression program of normal and amyloidogenic plasma cells.**

Please see the attached file in Source Data.

**Table S3. De-identified clinical characteristics of patients in the bulk RNA-seq cohort.**

Please see the attached file in Source Data.

**Table S4. Probability score of plasma cell-centered cell-cell communication in each sample.**

Please see the attached file in Source Data.

**Table S5. Source data and patient information of the multiplex immunofluorescence experiment.**

Please see the attached file in Source Data.

**Table S6. Antibodies used in multiplex immunofluorescence staining of bone marrow samples.**

Please see the attached file in Source Data.

**Table S7. Source data and patient information of the flow cytometry experiment.**

Please see the attached file in Source Data.
